# SARS-CoV-2 multi-variant graphene biosensor based on engineered dimeric ACE2 receptor

**DOI:** 10.1101/2021.10.02.21264210

**Authors:** Mattia D’Agostino, Eleonora Pavoni, Alice Romagnoli, Chiara Ardiccioni, Stefano Motta, Paolo Crippa, Giorgio Biagetti, Valentina Notarstefano, Simone Barocci, Brianna K. Costabile, Gabriele Colasurdo, Sara Caucci, Davide Mencarelli, Claudio Turchetti, Marco Farina, Luca Pierantoni, Anna La Teana, Richard Al Hadi, Mauro Chinappi, Emiliano Trucchi, Filippo Mancia, Blasco Morozzo della Rocca, Ilda D’Annessa, Daniele Di Marino

## Abstract

Fast, reliable and point-of-care systems to detect the SARS-CoV-2 infection are crucial to contain viral spreading and to adopt timely clinical treatments. Many of the rapid detection tests currently in use are based on antibodies that bind viral proteins^1^. However, newly appearing virus variants accumulate mutations in their RNA sequence and produce proteins, such as Spike, that may show reduced binding affinity to these diagnostic antibodies, resulting in less reliable tests and in the need for continuous update of the sensing systems^2^. Here we propose a graphene field-effect transistor (gFET) biosensor which exploits the key interaction between the Spike protein and the human ACE2 receptor. This interaction is one of the determinants of host infections and indeed recently evolved Spike variants were shown to increase affinity for ACE2 receptor^3^. Through extensive computational analyses we show that a chimeric ACE2-Fc construct mimics the ACE2 dimer, normally present on host cells membranes, better than its soluble truncated form. We demonstrate that ACE2-Fc functionalized gFET is effective for *in vitro* detection of Spike and outperforms the same chip functionalized with either a diagnostic antibody or the soluble ACE2. Our sensor is implemented in a portable, wireless, point-of-care device and successfully detected both alpha and gamma virus variants in patient’s clinical samples. As incomplete immunization, due to vaccine roll-out, may offer new selective grounds for antibody-escaping virus variants^4^, our biosensor opens to a class of highly sensitive, rapid and variant-robust SARS-CoV-2 detection systems.

---

Thanks to its high rate of human-to-human transmission, SARS-CoV-2 has rapidly become a health emergency of worldwide concern and magnitude, greatly impacting human lives and global economics^5–7^. To monitor the spread of SARS-CoV-2, several diagnostic tests are being developed^8,9^. The most widely employed are based on viral RNA amplification (molecular tests) or viral protein detection via specific antibodies (antigenic test)^10^. The former represents the gold standard among SARS-CoV-2 tests, but it requires a few hours and specialized machineries to be completed. Conversely, most antigenic tests are rapid but show poor sensitivity as a drawback^11–13^. In addition, some of the recently emerged SARS-CoV-2 variants, bear mutations in the standard targets of antigenic tests (*i*.*e*., viral Spike and Nucleocapsid protein), potentially impacting the ability of antigenic tests to specifically recognize the virus^14–16^. Virus variants^17^ will keep appearing as long as the pandemic is not contained and an incompletely immunized host population (*i*.*e*., due to slow vaccine roll-out, with delays between the two doses, or because of declining protection a few months after the complete vaccination) may favour selection of antibody-escaping virus variants^4^. Thus, alternative variant-robust biosensors, capable of rapidly detecting SARS-CoV-2, have vital importance in keeping the COVID-19 outbreaks under control. Thanks to their sensitivity and rapidity, graphene field-effect transistors (gFET)^18^, recently proposed also for virus detection^19,20^, represent a promising biosensing approach. In a gFET, a graphene monolayer connects the source and drain electrodes of a transistor and the graphene is functionalized with a bioreceptor able to specifically bind target molecules. The bioreceptor-target interaction alters graphene’s electronic properties resulting in a readily detectable signal^18^. gFETs are thus attractive in point-of-care diagnosis due to their miniaturization, the potential for large-scale manufacture and operability by non-specialized personnel.

Here, we developed a gFET biosensor that uses ACE2 as bioreceptor, aiming to mimic the viral mechanism of host cell access, whereby the viral Spike protein recognizes and binds ACE2, a widely expressed membrane-anchored protein^21^ (Fig. 1A). Spike is a trimeric transmembrane protein whose monomers are composed by two subunits, S1 and S2, with S1 containing the receptor-binding domain, RBD (Fig. 1B)^22^. RBD in the “up” conformation is competent for binding^23^. ACE2 (Fig. 1C) is a widely expressed transmembrane protein composed by a collectrin-like domain (CLD) that ends with a single transmembrane helix and by a peptidase domain (PD), the latter being specifically bound by RBD^23^. Contrarily to antibodies (Ab) elicited by a former virus variant which can be eluded by newly appearing variants (immune escape)^24^, the key contact point established by the virus Spike and the host ACE2 conditions the host cell infection and showed increased affinity in some of the most rapidly spreading variants^3^. Therefore, ACE2-Spike appears to be a natural bioreceptor-target pair for variant-robust SARS-CoV-2 sensing.

**Fig. 1.**
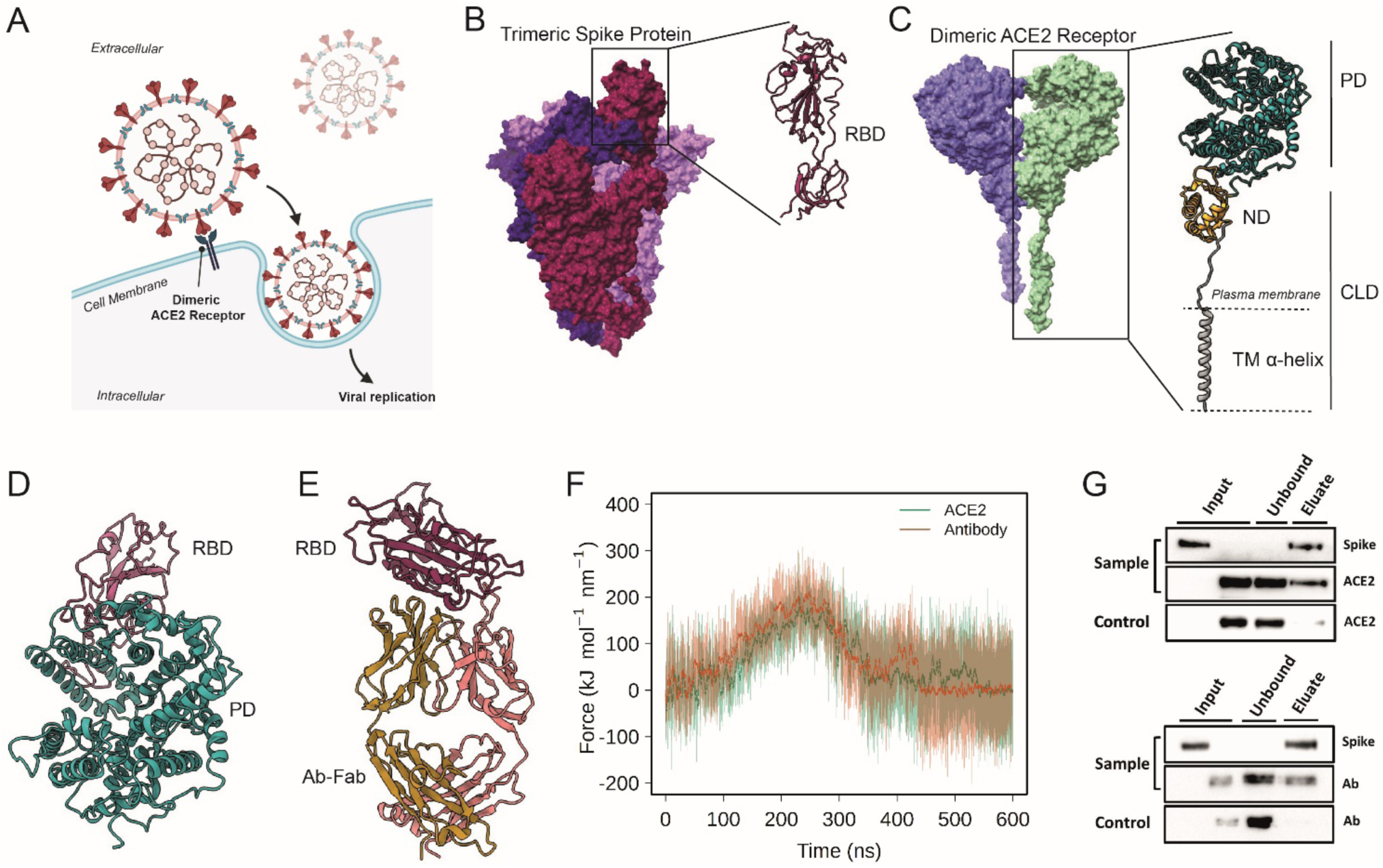
Probing the interactions of ACE2 and Antibodies with Spike. (**A**) Schematic representation of ACE2-mediated host cell entry mechanism. (**B**) Cryo-EM structure of soluble trimeric Spike protein (the three monomers have different colors). A zoomed in view of the RBD is shown. (**C**) Cryo-EM structure of dimeric ACE2 receptor (the two monomers are colored differently). The two subunits of a monomer are reported: Peptidase domain (PD) and Collectrin-like domain (CLD) that is composed by neck domain (ND) and transmembrane helix (TM). (**D**) Peptidase Domain of the ACE2 receptor bound to the RBD of the SARS-CoV-2 Spike protein. (**E**) Antibody CR3022, bound to the SARS-CoV-2 spike protein RBD. (**F**) Force profiles from the steered MD simulations of RBD unbinding from the ACE2 receptor (orange) and Ab-CR3022 (green) (**G**) Pull-down assay of Spike and ACE2 (upper), and Spike and Ab Anti-Spike (lower). Control is represented by the same experiment excluding the Spike protein (bait) from the system. The binding of Spike with ACE2 or anti-Spike were monitored by Western blot analysis.

## Soluble ACE2 and Ab binding with Spike-RBD

As a first step in exploring the possibility of using ACE2 as a bioreceptor, by performing a series of constant velocity Steered Molecular Dynamics (SMD) simulations, we characterized in silico the ACE2-RBD complex (Fig. 1D) in comparison with an Ab-RBD complex (Fig. 1E). Since RBD is the interaction domain^23^, this was used as a proxy for Spike in our simulations. To account for the various known RBD binding sites we used as initial starting point for SMD three representative structures obtained by structural comparison (Extended Data Figure 1). Although they show different patterns of interaction, the force needed to dissociate the RBD from ACE2 or from the most stable Ab-RBD complex (group 3) is the same (Fig. 1F) at both velocities tested (Extended Data Figure 2). A detailed description of the ACE2-RBD unbinding mechanism is in Extended Figure 3. The other two groups exhibited lower binding forces and were thus less relevant in the comparison.

The interaction of ACE2, in its soluble truncated form, with the trimeric Spike protein was also probed with pull-down assays and western blot (Fig. 1G). Spike-decorated sepharose beads were used to pull down ACE2 and both are recovered in the eluted fractions, confirming their specific interactions. No Spike leakage was observed in the unbound fractions on each pull-down assay (Fig. 1G, unbound lanes). Similar pull-down results were obtained for the anti-Spike Ab (CR3022). Taken together, our biochemical and computational results suggest that ACE2 can be used as a bioreceptor for gFETs functionalization.

## Spike detection by ACE2_gFET and Ab-CR3022_gFET

Spike detection was performed using a gFET provided by Graphenea (San Sebastian, Spain) consisting of 12 separated single-layer graphene channels connected to source and drain gold electrodes. A non-encapsulated electrode at the centre of the chip allows liquid gating through PBS solution (Fig. 2A). The bifunctional 1-pyrenebutanoic acid succinimidyl ester (PBASE) was used as a linker between graphene and bioreceptor proteins (ACE2 and Ab-CR3022). PBASE pyrene group stacks via π-π interactions on the graphene aromatic lattice, while the succinimide covalently binds amino groups of proteins^25^. To obtain efficient coverage, avoiding the formation of multiple layers of pyrene^26^, we treated the gate electrode with a 5 mM PBASE in DMF solution. We characterized the bare and functionalized gFET by Atomic Force Microscopy (AFM) (Extended Data Figure 4A-D) and Raman spectra^27^ (Fig. 2B). Electrical characterization of pristine and activated gFET further confirms the PBASE attachment to graphene (Extended Data Figure 5).

**Fig. 2.**
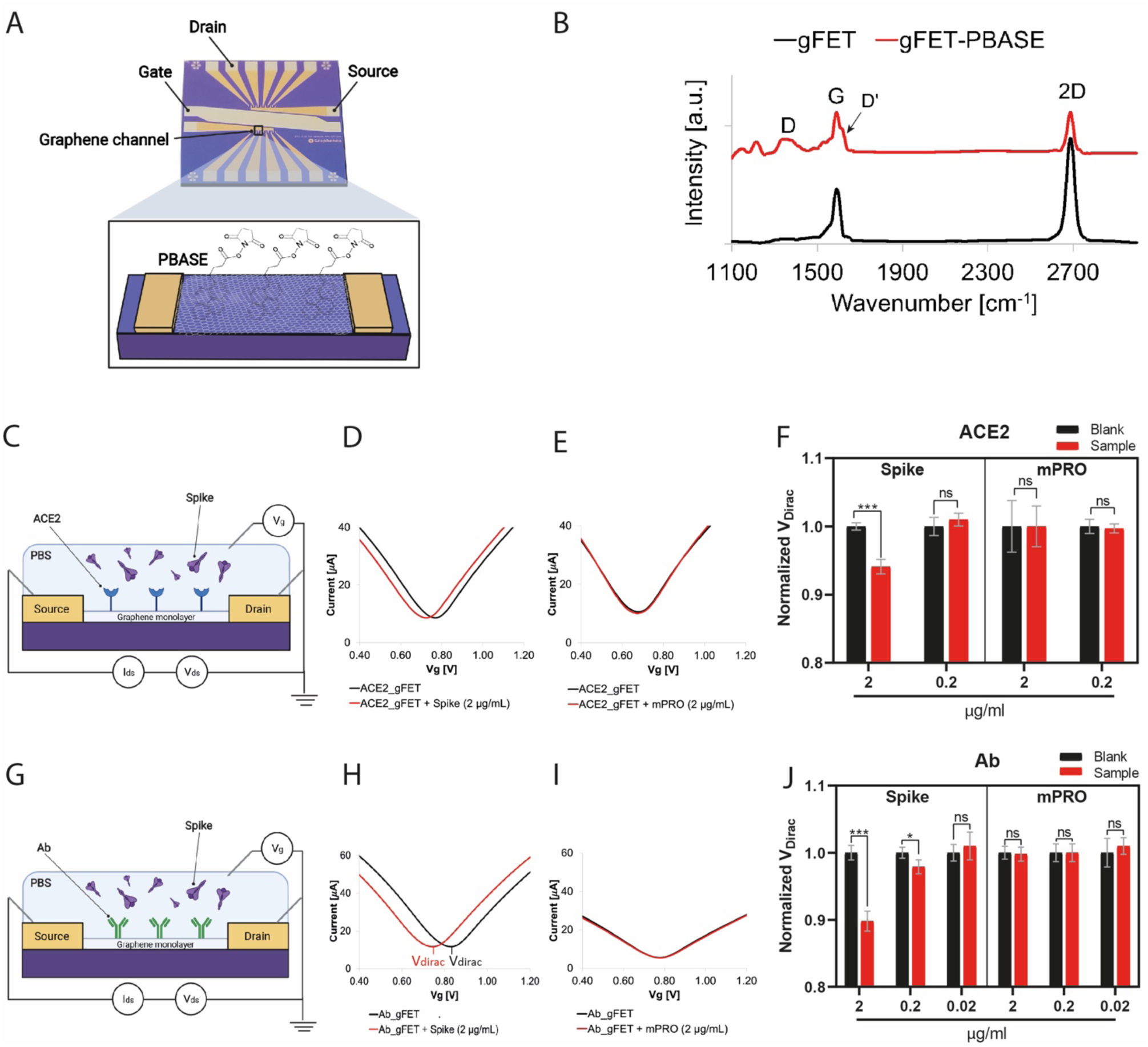
gFET setup and Spike recognition. (**A**) gFET (size 10 mm × 10 mm) is composed by two source electrodes each one connected with six graphene channels and the respective drains. A single gate electrode is used for both sides of gFET. A schematic representation of the PBASE-modified gFET is reported in the inset panel. (**B**) Raman Spectra of gFET (black) and gFET-PBASE (red) (diode laser wavelength 523 nm and laser power 50 mW). (**C**) Schematic representation of gFET modified with ACE2. (**D-E**) I_ds_-V_g_ curves obtained as the mean of six different measurements on the same device of **(D)** ACE2_gFET (black) and ACE2_gFET-Spike (red), Spike protein was 2 µg/mL. (**E**) ACE2_gFET (black) and ACE2_gFET-mPRO (red). mPRO protein was 2 µg/mL. (**F**) Comparative bar charts of ACE2_gFET before (black bars) and after (red bars) the addition of different concentrations of Spike or mPRO. Data were normalized on V_dirac_ value max for each measurement average. (***P<.001, mean n=6, n.s. not significant P>.05, t-test discovery determined using the Two-stage linear step-up procedure of Benjamini, Krieger and Yekutieli, with Q = 1%. Error bars represent s.d.). (**G**) Schematic representation of gFET modified with Ab. (**H-I**) I_ds_-V_g_ curves obtained as the mean of six different measurements on the same device of **(H)** Ab_gFET (black) and Ab_gFET-Spike (red), Spike protein was 2 µg/mL, V_dirac_ is marked; (**I**) Ab_gFET (black) and Ab_gFET-mPRO (red), mPRO protein was 2 µg/mL; (**J**) Comparative bar charts of Ab_gFET before (black bars) and after (red bars) the addition of different concentrations of Spike or mPRO. Data were normalized on V_dirac_ value max for each measurement average. (***P<.001, mean n=6, n.s. not significant P>.05, t-test discovery determined using the Two-stage linear step-up procedure of Benjamini, Krieger and Yekutieli, with Q = 1%. Error bars represent s.d.).

Next, we investigated the sensing performances of the gFET functionalized with the soluble ACE2 (Fig. 2C-F) and antibody Ab-CR3022 (Fig. 2G-J). Experiments were conducted fixing the drain-source voltage to V_ds_=50 mV and varying the gate voltage V_g_ from 0 to 1.5 V. The transfer curves present a minimum (Dirac point) whose position is altered when the electronic structure of graphene is perturbed, *i*.*e*., by the bioreceptor-target interaction^18^. Decreasing Spike protein concentrations were serially tested. For ACE2_gFET after the addition of the Spike protein solution at 2 μg/mL, a negative shift of transfer curves is observed (Fig. 2D). Applying a Spike concentration of 0.2 μg/mL instead, resulted in no significant differences. When we tested the Ab_gFET, a negative shift was observed both at 2 and 0.2 μg/ml. Subsequent dilution (0.02 μg/ml) did not elicit a detectable Dirac point shift (Extended Data Figure 6). Furthermore, the specificity of the bioreceptor-target binding is supported by the absence of significant shifts observed when using mPRO, the main protease of the same virus, (Fig 2E, 2I and Extended Data Figure 6) at all concentrations.

## ACE2-Fc chimera mimics the transmembrane ACE2 dimer

To overcome the lower sensitivity of soluble truncated ACE2_gFET as compared to Ab_gFET, we implemented a chimeric version of ACE2 fused with a Fc-tag at its C-terminus (ACE2_Fc). We expected that the disulfide bridges present in the Fc-tag^28^ would enforce the formation and stability of dimeric ACE2 complex, mimicking what occurs in physiological conditions on the cell membrane^23^.

We computationally characterized via MD simulations three systems: i) a complete ACE2 system with transmembrane helices embedded in a POPC:CHOL (90:10) membrane (Fig. 3A-B); ii) a chimeric system composed of the soluble portion (PD, CLD, ND) of ACE2, linked to Fc (Fig 3D), and the iii) the soluble ACE2 form (Fig. 3C). In the ACE2-Fc chimeric system, after a transient period of about 100 ns, we observed a stable dimeric conformation for the rest of the 500 ns sampled (Fig. 3F). This dimeric system is similar to the membrane embedded one. The distance between the PD domains centre of mass was used to monitor intramonomer distance (Fig. 3E). In both membrane embedded ACE2 and chimeric ACE2-Fc systems distance fluctuates around a value of 7 nm, rarely sampling more open conformations (of up to 8 nm). On the contrary, in soluble ACE2 the monomers tend to separate more (Fig. 3G grey curve). The rearrangement of the monomers is also evident from the time evolution of the number of contacts between monomers involving the PD and CLD regions, with the soluble ACE2 system losing more contacts over time (Fig. 3H) and from the comparison of the RMSD of the two PD domains (Extended Data Figure 7A). Overall, these results indicate that the chimeric ACE2-Fc system better preserves the dynamical properties of ACE2 in membranes (Supplementary Video 1). This was made possible by an overstabilization of the CLD domains due to the closer distance of the CLD-Fc connecting linker (Extended Data Figure 7B) that are held together by the disulfide bridges within the Fc region (Extended Data Figure 8).

**Fig. 3.**
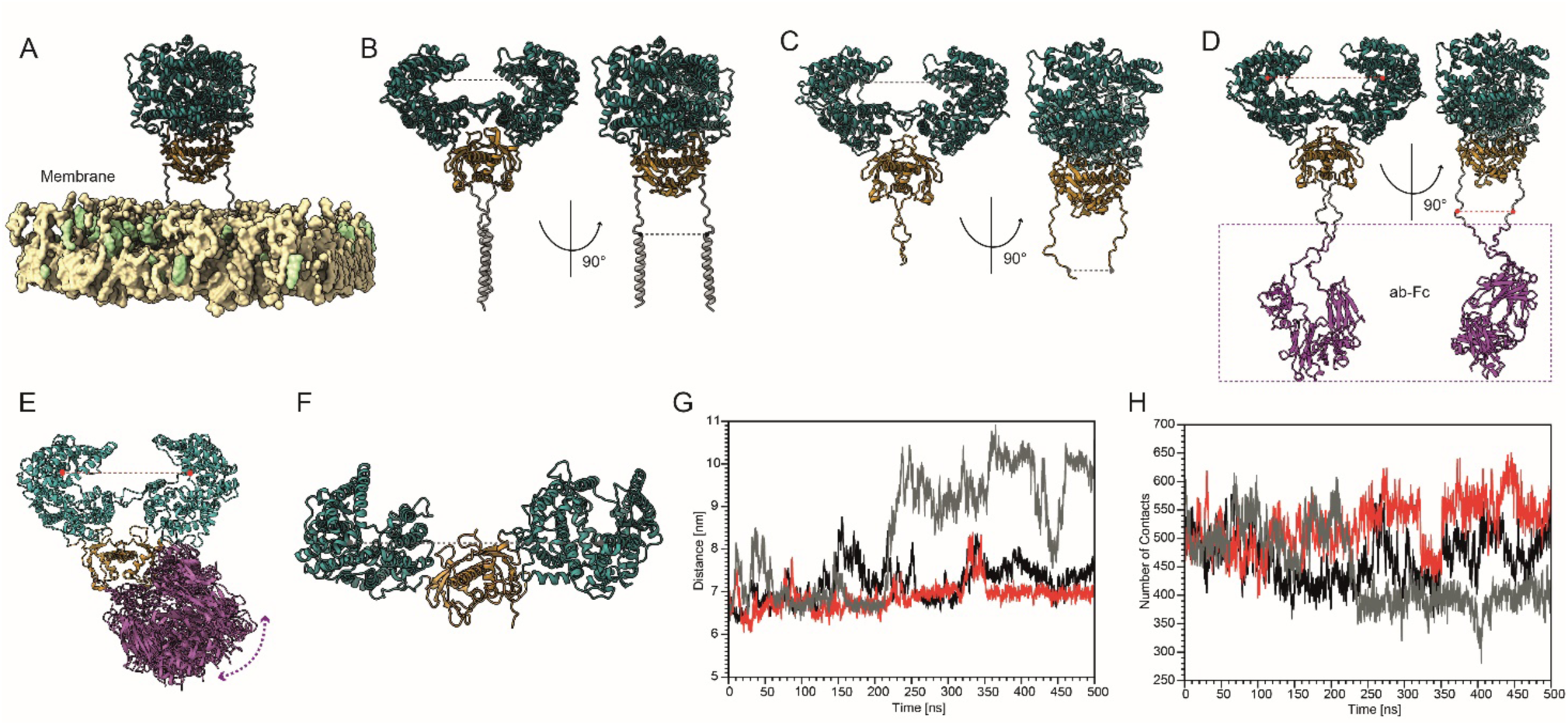
MD simulations of full length, soluble and Fc tagged ACE2 dimers. (**A**) Representative structure of the full length ACE2 dimer embedded in a membrane. (**B**) Same as (**A**) but the membrane is omitted to show the TM helices. (**C**) Soluble ACE2 conformation. (**D**) Starting configuration of the ACE2-Fc chimera. For **B**-**D** two orthogonal views are shown. (**E**) Representative snapshots of ACE2-Fc structures sampled during the MD trajectory, side view. The PD centres of mass distance is shown by a dashed red line (**F**) top view of soluble ACE2, the PD centres of mass distance is shown by a dashed black line. (**G**) Time evolution of the intermonomer distance measured between the PD domains for membrane embedded full length ACE2 (in black), ACE2-Fc (red) and soluble ACE2 (grey). (**H**) Number of contacts between the two PD-CLD regions of monomers, color code as in (**G**).

The dimerization propensity of the ACE2-Fc chimera inferred from computational investigation was confirmed by performing SDS-PAGE electrophoresis with (Wβ) and without (W/o β) β-mercaptethanol. When ACE2-Fc is in reducing conditions (Fig. 4A, Wβ), the disulfide bonds in the Fc region are reduced and the protein migrates as a monomer. On the other hand, ACE2-Fc in non-reducing condition runs as a dimer (Fig. 4A, W/o β), consistently with the computational results. Soluble ACE2, as expected by the absence of disulfide bonds, is not affected by reducing conditions. The interaction of ACE2-Fc with the trimeric Spike protein was also confirmed with pull-down assays and western blot (Fig. 4B).

**Fig. 4.**
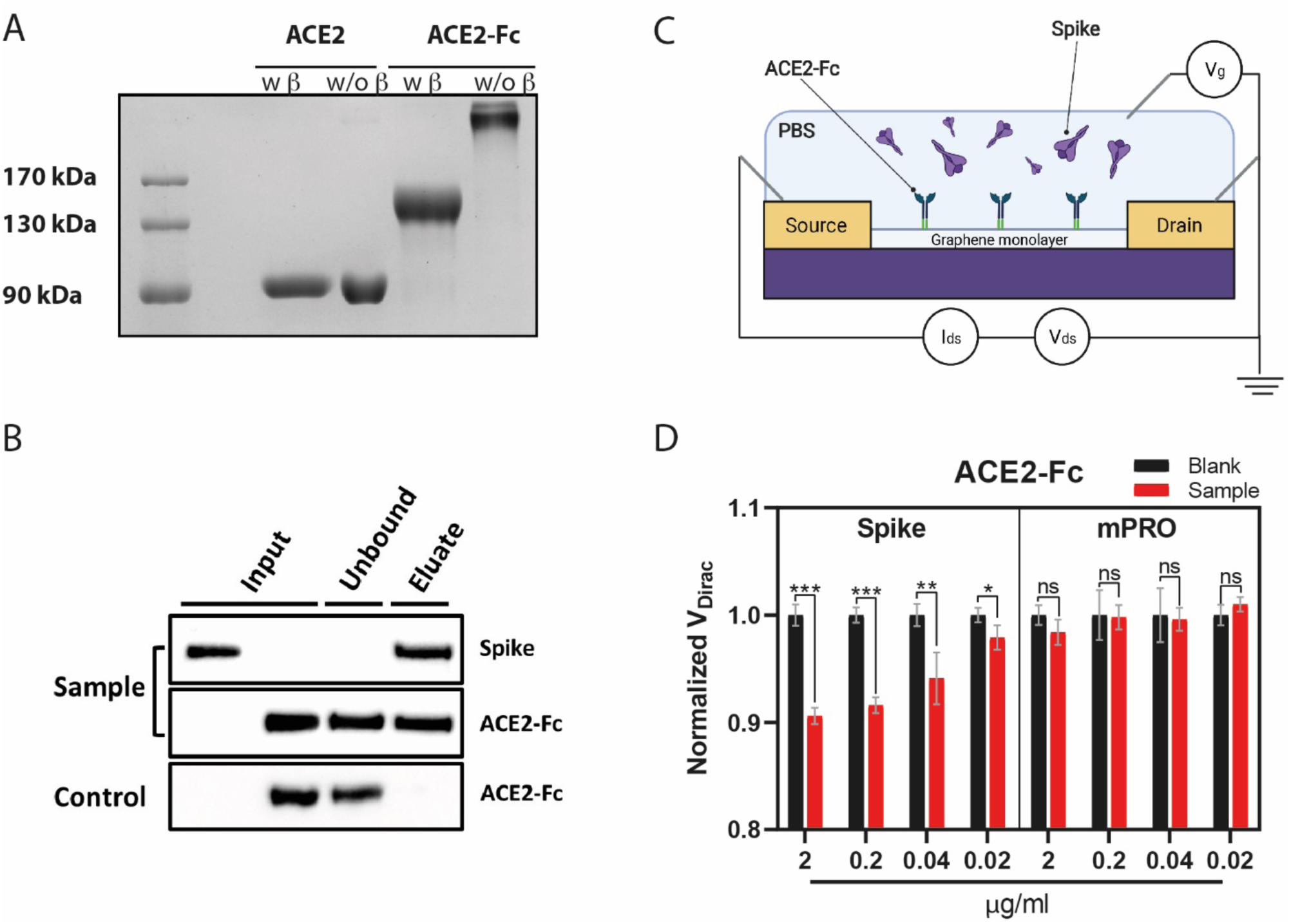
ACE2-Fc as bioreceptor. (**A**) SDS-PAGE under reducing (w β lanes) and non-reducing (w/o β lanes) conditions of soluble ACE2 and ACE2-Fc; (**B**) Pull-down assay of Spike and ACE2-Fc. The binding of Spike with ACE2-Fc was monitored by Western blot. (**C**) Schematic representation of gFET modified with ACE2-Fc. (**D**) Comparative bar chart showing the ACE2-Fc_gFET response to different concentrations of Spike (left) or mPRO (right); ***P<.001, **P<.01 and *P<.05.

## Spike detection by ACE2-Fc_gFET

A gFET was functionalized with ACE2-Fc (Fig. 4C) and tested using different Spike concentrations (Fig. 4D and Extended Data Figure 9). Even at the lowest concentration used, 0.02 μg/mL, the ACE2-Fc_gFET is able to detect the Spike protein (Extended Data Figure 9). No significant differences are observed for mPRO (Fig. 4D). Thus, the dimeric ACE2-Fc bioreceptor is specific for Spike and outperforms both the soluble ACE2 (no signal for 0.2 ug/ml, Fig. 2F) and the Ab-based system (no signal for 0.02 ug/ml, Fig. 2J). Notably our sensitivity is comparable to that of lateral flow based devices^29^. Since ACE2 is the receptor for only three coronaviruses, one of which is contained and the other, HCoV-NL63, has low binding affinity, specificity is reasonably assured^30^.

## Detection of SARS-CoV-2 from swab by point-of-care device

To achieve portability, we designed and built a customized reusable point-of-care (POC) device (Fig. 5A-B) accommodating the gFET chip and giving the same readouts of the lab scale probe station used during sensor implementation and testing (Extended Data Figure 11). The manufacturing process is described in the Supplementary Notes 4 and 5, as well as the solutions adopted to realize an energy-efficient, battery-powered, Bluetooth device able to reliably detect currents around 10 μA and Dirac voltage shifts of a few millivolts. Nasopharyngeal swab specimens from three different patients, stored in PBS 1X, have been analysed with ACE2-Fc_gFET (Fig. 5C) loaded on our POC device. RT-qPCR results indicate that two patients were positive to SARS-CoV-2, one carrying the alpha and the other the gamma variant, while the third tested negative (Fig. 5E). The gFET transfer curves are reported in Extended Data Figure 11, while the normalized Dirac voltage is in Figure 5D, demonstrating that our POC device correctly identifies the positive patients. Interestingly, for these cases, the gFET signal shows the same qualitative trend of the RT-qPCR: higher viral load (see Ct average in Fig. 5E) corresponds to stronger gFET signal. Notably, no previous concentration/purification steps were executed on the patient samples. Wireless connectivity of the POC enables testing in a separate environment with respect to signal acquisition.

**Fig. 5.**
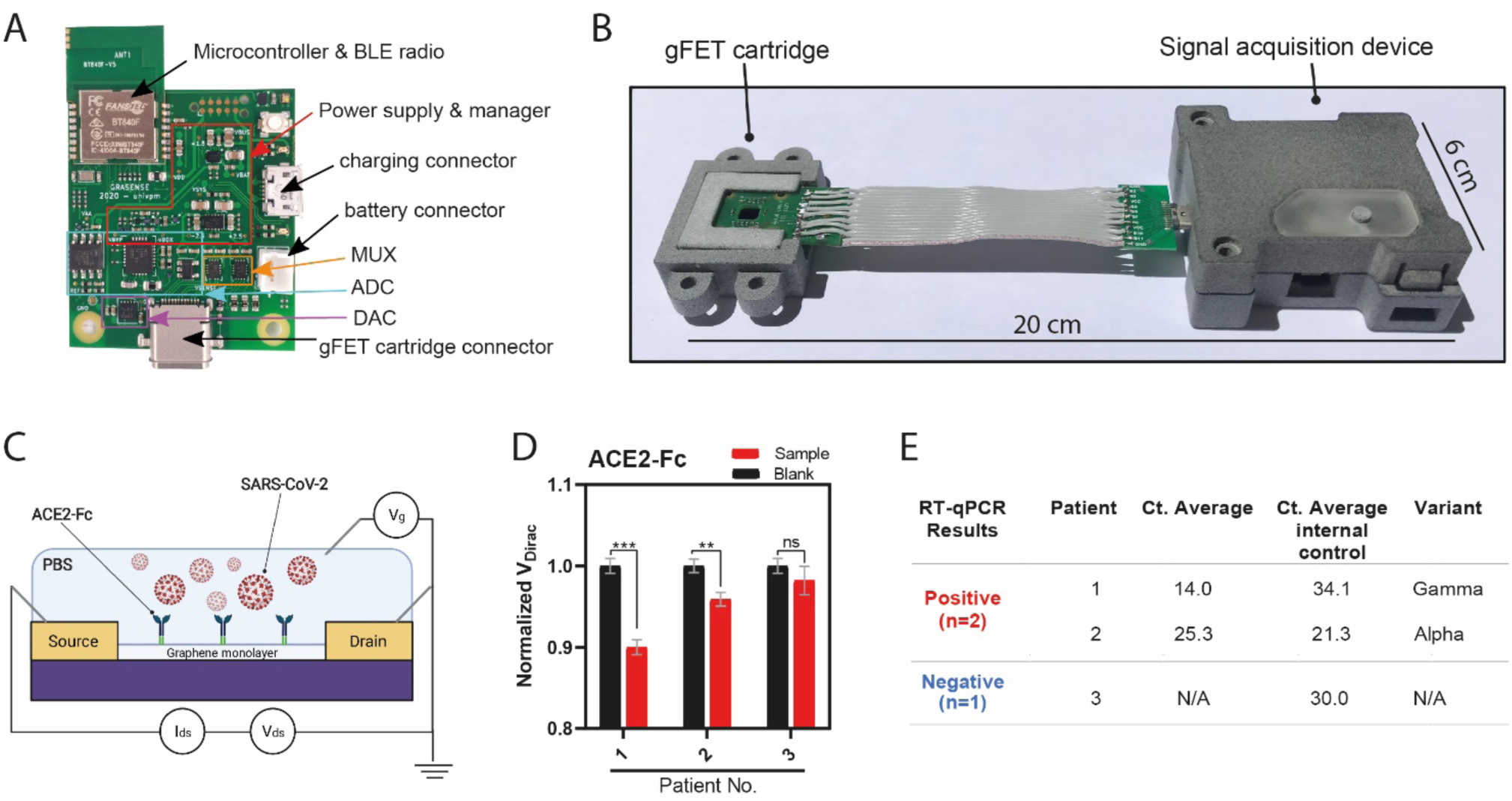
Point-of-Care (POC) device detects SARS-CoV-2 in patient samples. (**A**) Signal acquisition module of the POC device with main elements labelled. (**B**) Photograph of the gFET Cartridge Unit and the Signal acquisition modules connected together to form the entire POC. A reference dimension bar is reported below. (**C**) Schematic representation of gFET modified with ACE2-Fc tested on viral samples from patients. (**D**) Bar graph reporting ACE2-Fc_gFET signal before (black) and after the addition of swab specimens (red) from patient 1, 2 and 3. **P<.01, ***P<.001; (**E**) RT-qPCR results for the detection of the SARS-CoV-2 specific genes (E gene; RdRP gene; S gene and ORF1ab gene) of the three patient samples. Ct value in clinical samples were evaluated. Ct average value of internal control are indicated for each sample. The type of virus variant (as detected by targeted RT-PCR) is also reported.

## Discussion

Rapid, sensitive and variants-robust detection systems have been immediately acknowledged as crucial in the COVID-19 global containment strategy. Today, as the global society urgently needs to reopen to people interactions and circulation, while vaccines are unevenly rolling out throughout the world and the virus is far from being contained, accurate detection tests able to provide quick responses are even more necessary. Highly sensitive PCR-based molecular tests still require a few hours for the result, lab scale facilities and specialized personnel, whereas quick antigen-based tests are characterized by lower accuracy^13^, which is expected to be further affected by newly emerging virus variants^31^. Moreover, both testing approaches employ disposable plastic supplies in their procedures raising concerns about the environmental impacts associated with the global scale testing campaigns^32^.

Here we present a gFET sensor that exploits as bioreceptor a chimeric version of the human receptor ACE2 which outperforms an anti-Spike antibody (Ab-CR3022) in detecting the viral Spike protein. The fusion of the Fc-tag on ACE2’s C-terminus promotes stable protein dimerization, making it more similar to its native state on cell membrane^28^ as shown by our computational and *in vitro* biochemical analyses. Mimicking the actual virus-host interaction for cell infection, whose affinity has been shown to have improved in some of the late variants^3^, our sensor is expected to be robust to current and future virus variants.

Our biosensor, miniaturized into a reusable, low-waste user-friendly POC device, was successfully tested with clinical samples from patients infected by alpha and gamma virus variants. Accuracy studies coupled with PCR-based molecular tests as a benchmark will be, of course, needed to verify the performance of our POC biosensor with different and upcoming virus variants. In addition, modifications of the ACE2 amino acid sequence, which are expected to increase binding affinity with the viral Spike^33^, could also further improve the sensitivity of our ACE2-Fc gFET-based biosensor. In general, our novel biosensor set the basis for a class of highly-sensitive, fast and variant-robust SARS-CoV-2 detection systems.

## Data Availability

Relevant data are available from the corresponding author upon reasonable request.

## Methods

### Ethics approval

All subject provided written consent to be used for research purposes.

The Ethics Committee of Region Marche (CERM) approved the study on 17/12/20. Components of CREM are listed here https://www.ospedaliriuniti.marche.it/portale/index.php?id_sezione=132&id_doc=446&sottosezione=37

### Structural Clustering of RBD binding modes in experimental complexes

The structural comparison among all available complexes of RBD with Antibody or ACE2 (see Supplementary note 1) revealed 3 different binding modes, shown in Extended Data Figure 1. We choose a representative structure of each group to simulate (PDBID: 7BEK, 7MF1 and 6YLA) and compared the SMD forces necessary to obtain a complete unbinding.

### Steered Molecular Dynamics simulations

Structures were first equilibrated with a multistage equilibration protocol adapted from^34^ (details in Supplementary note 2). The SMD simulations were performed by harmonically restraining the x component of the distance between the centre of mass of the backbone of the two proteins with a force constant of 10 kJ mol^-1^ Å^-2^. Two different simulations for each system were performed with a constant velocity of 0.05 Å ns^-1^ or 0.02 Å ns^-1^. See Supplementary note 2 for details.

### Simulation of dimeric ACE2

The three systems (ACE2-Membrane, soluble ACE2 and ACE2-Fc) were built starting from the structure 6M17. Systems were properly equilibrated with a multistage equilibration protocol (Supplementary note 3) and then simulated in absence of restraints. The first 30 ns of each simulation was discarded as a further equilibration stage, and the subsequent 500 ns were analysed.

### Non-reducing SDS-PAGE

The ACE2-Fc dimerization was assessed through SDS-PAGE^35^ and carried out under non-reducing and reducing conditions. Briefly, 2µg of ACE2-Fc or ACE2 samples were placed 10 minutes at 100 °C under denaturing conditions with Laemmli sample buffer reduced by β-mercaptoethanol or under non-reducing condition using a sample buffer without β-mercaptoethanol. 8% gel was used to correctly separate ACE2-Fc or ACE2 monomers from the dimers.

### Purification of Trimeric Spike protein from FreeStyle HEK293-F cells

The plasmid for expression of the SARS-CoV-2 prefusion-stabilized Spike ectodomain in HEK293-F cells (Thermo Fisher) was a generous gift from the McLellan laboratory at the University of Texas at Austin^36^. 350 μg and 1.05 mg PEI (Polysciences Inc.) were used to transfect cells at 1.2×10^6^ cells/ml^37^. After five days in suspension culture, the cell supernatant was collected and filtered using a 0.22 μm filter. Protein in the supernatant was bound to Nickel-NTA agarose (Qiagen) while under rotation in buffer composed of 2 mM Tris, 150 mM NaCl and 10 mM imidazole for two hours at 4°C. The resin was then washed in Tris-NaCl buffer, pH 8, with 20 mM imidazole and protein was eluted in 200 mM imidazole. Following overnight dialysis against PBS, the protein was filtered and stored as 0.2 mg/ml aliquots at -80°C.

### Pull-down assay

The protein pull-down assay to validate the interaction between Spike and ACE2 or anti-Spike antibody Ab-CR3022 or ACE2-Fc was performed using a strep-tactin Sepharose resin as previously described^38^, with some modifications. Briefly, 80 μl indicated strep-tactin resin were washed with phosphate-buffered saline pH 7.4 (PBS 173 mM NaCl, 2.7 mM KCl, 10 mM Na_2_HPO_4_, 2mM KH_2_PO_4_) and incubated with 5 μg of recombinant trimeric Spike protein with a C-terminus strep-tag (Spike-strep) next to an his-tag at room temperature for 1 hour. After incubation, Spike-bound beads were washed three times with 500 µl PBST buffer (PBS and 0.05% Tween-20) and then were aliquoted into different tubes. 5 μg of ACE2 in PBS or 5 μg of anti-Spike or 5 μg of ACE2-Fc in PBS were mixed with spike-bound beads in three different 1.5 ml tubes and incubated at room temperature for 1 hour separately. After a 1-hour incubation, beads were washed three times with 500 µl PBST buffer and the bound proteins were eluted using 50 µl of elution buffer (PBS added of 2.5 mM biotin (Sigma)). The samples were then subjected to SDS–PAGE and analysed by western blotting using an anti-Histidine antibody (Thermo Scientific) to detect Spike and ACE2, an anti-rabbit antibody (Santa Cruz Biotechnology) to detect Ab-CR3022 and an anti-ACE2 antibody (EMP Millipore Corp) to detect ACE2-Fc by chemiluminescent revelation^39^. The same protocol, using empty beads (without Spike), was performed as a negative control for each system.

### gFET functionalization

ACE-Fc, ACE2 and anti-Spike antibody were covalently immobilized over the fabricated gFET chip (Graphenea gFET-S20) through PBASE. A droplet of 5 mM PBASE (Thermo Fisher Scientific, Waltham, MA) in dimethylformamide (DMF) was placed on the chip for 2h at room temperature before being rinsed several times with DMF, deionized water (DI) and dried with N2. Finally, the PBASE-functionalized devices were exposed to 250 μg/mL of ACE2-Fc (Z03484-1; GenScript Biotech), ACE2 (10108-H08B-100; Sino Biological, Inc., China) or anti-Spike (40150-R007; Sino Biological, Inc., China) separately and left overnight in a humidified environment at 4 °C. The sensor was then sequentially rinsed in PBS (pH 7.4, 1X), DI water and dried under N2 flow. The chip was subsequently treated with 100 mM glycine in PBS (pH 8.4, 1X) for 30 minutes for the termination of excess PBASE NHS groups at room temperature. After glycine treatment, samples were rinsed with PBS (pH 7.4, 1X), DI water and dried with N2.

### gFET characterization using Raman and AFM

AFM measurements were performed with a SOLVER PRO from NT-MDT, RMS was evaluated by using Nova Px software. A Horiba Jobin-Yvon XploRA Raman microspectrometer, equipped with a 532-nm diode laser (∼50mW laser power at the sample) was used. All measurements were acquired by using a ×100 long working distance objective (LMPLFLN, N.A. 0.8, Olympus). The spectrometer was calibrated to the 520.7 cm^-1^ line of silicon prior to spectral acquisition. A 2400 lines per mm grating was chosen. The spectra were dispersed onto a 16-bit dynamic range Peltier cooled CCD detector. The spectral range from 1100 to 3000 cm^-1^ was chosen and spectra were acquired for 3×10 seconds at each measurement spot. Chips were measured before and after PBASE functionalisation. For each sample, 10 point/spectra were acquired, and a Raman map was acquired with the same parameters on squared areas (20 µm × 20 µm), with a step size of ∼3 µm, for a total number of 36 spectra. On each Raman map, the following values were calculated: intensity of the band centred at 2690 cm^-1^ (the 2D band), the intensity of the band at 1592 cm^-1^ (the G band), and the ratio between these two bands (I_2D_/I_G_). False colour images were built by using the I_2D_/I_G_ ratio.

### gFET electrical measurements

Sensing performances were evaluated by using a Wentworth probe station equipped with a Bausch & Lomb MicroZoom optical microscope and by using an HP4145B semiconductor parameter analyser. Current-voltage curves (I_ds_-V_ds_) have been acquired (i) by applying a V_ds_ between –0.1 V to 0.1 V, (ii) by operating in liquid gating condition with PBS solution pH 7.4, and (iii) by using a Vg of 0 V. Transfer curves (I_ds_-V_g_) have been obtained (i) by using V_ds_ 0.050 V, (ii) by operating in liquid gating condition with PBS solution at pH 7.4, (iii) by applying Vg swept from 0 to 1.5 V, (iv) by carrying out a relaxation step to obtain a constant equilibrium of ions on the surface of graphene^19^. During this step, V_ds_ and V_g_ were both applied on the gFET until no variations on the current-voltage curves were observed; in this way, the same ions screening effect was maintained during the measurements and the current-voltage curves, taken on the same gFET at different times, were completely superimposable. Recombinant mPRO, used as a negative control, was kindly provided by Prof. Paolo Mariani from Polytechnic University of Marche^40^.

### RT-qPCR of patient samples

Clinical Samples used in this study (Table 1) were kindly provided by Dott. Simone Barocci from U.O.C. of Clinical Pathology from the hospital of Urbino (Italy) “Santa Maria della Misericordia”. Nasopharyngeal swabs from COVID-19 positive patients and COVID-19 negative were stored in PBS 1X and used. The positivity or negativity of these samples were determined by real-time RT-qPCR following manufacturer’s specifications (ALLPLEX SARS-CoV-2 ASSAY and MDS methodologies).

### RT-PCR SARS-CoV-2 and Variants Detection

Viral RNA was extracted from nasopharyngeal swabs using the Kit QIAsymphony DSP Virus/Pathogen Midi kit on the QIAsymphony automated platform (QIAGEN, Hilden, Germany) according to manufacturer’s instructions. Multiplex real-time RT-PCR assay was performed using qPCRBIO Probe 1-Step Go No-Rox (PCRBIOSYSTEMS, London, UK) on the Applied Biosystems 7500 Fast Dx Real-Time PCR Instrument (Thermo Fisher Scientific). The oligonucleotide primers and probes were designed to detect 69/70 deletion and N501Y mutation from virus spike gene to discriminate alpha and gamma lineage respectively. Variant lineage was confirmed by sequence analysis of spike gene using ABI Prism 3100 Genetic Analyzer (Applied Biosystems-HITACHI).

## Data availability

Relevant data are available from the corresponding author upon reasonable request.

## Acknowledgements

DDM acknowledges funding from FISR2020 program, grant number 03475 From the Italian Ministry of Research. We acknowledge the CINECA as part of the agreement with the University of Milano-Bicocca for the availability of high-performance computing resources.

## Author contributions

DDM conceived and designed the study. MDA, EP, AR and ALT designed the protocols and performed experiments on gFET. CA and PC contributed to electrical measurements. GB and CT designed the electronics of the device. VN performed Raman experiments. SM, DDM and IDA performed and analysed the MD simulations. EP performed AFM experiments. SB and SC contributed to patient samples collection and analysis. BKC and FM expressed the Spike protein. DM, MF, LP, MC, BMDR, and ET contributed to data interpretation. GC designed the case and carrier units of POC. RAH contributed to the design of the POC. SM, DDM, MC, EP, MDA and BMDR prepared the figures. DDM, ET, FM, ILD, MC, MDA, AR and BMDR wrote the manuscript. All authors reviewed the manuscript.

## Abbreviations

Ab: Antibody
Ab_gFET: Antibody functionalised graphene Field-Effect Transistor
ACE2: Angiotensin Converting Enzyme 2
ACE2-Fc: ACE2 chimeric construct with Fc at C-terminus
ACE2-Fc_gFET: ACE2-Fc functionalised graphene Field-Effect Transistor
ACE2_gFET: ACE2 functionalised graphene Field-Effect Transistor
AFM: Atomic Force Microscopy
CLD: Collectrin-like domain
Ct: Cycle Threshold
Fc: Fragment Crystallizable
gFET: graphene Field-Effect Transistor
MD: Molecular Dynamics
mPRO: main Protease of SARS-CoV-2
PBASE: 1-pyrenebutanoic acid succinimidyl ester
PBS: Phosphate buffered Saline
PCR: Polymerase Chain Reaction
PD: Peptidase Domain
POC: Point of Care
POPC:CHOL: palmitoyl-2-oleoyl-sn-glycero-3-phosphocholine Cholesterol
RBD: Receptor Binding Domain
RMSD: Root Mean Square Deviation
RT-qPCR: Real-Time quantitative Polymerase Chain Reaction
SARS-CoV-2: Severe Acute Respiratory Syndrome of Coronavirus 2
SDS-PAGE: Sodium Dodecyl Sulfate Polyacrylamide Gel Electrophoresis
SMD: Steered Molecular Dynamics
TM: Transmembrane

## Extended Data File

**Extended Data Figure 1.**
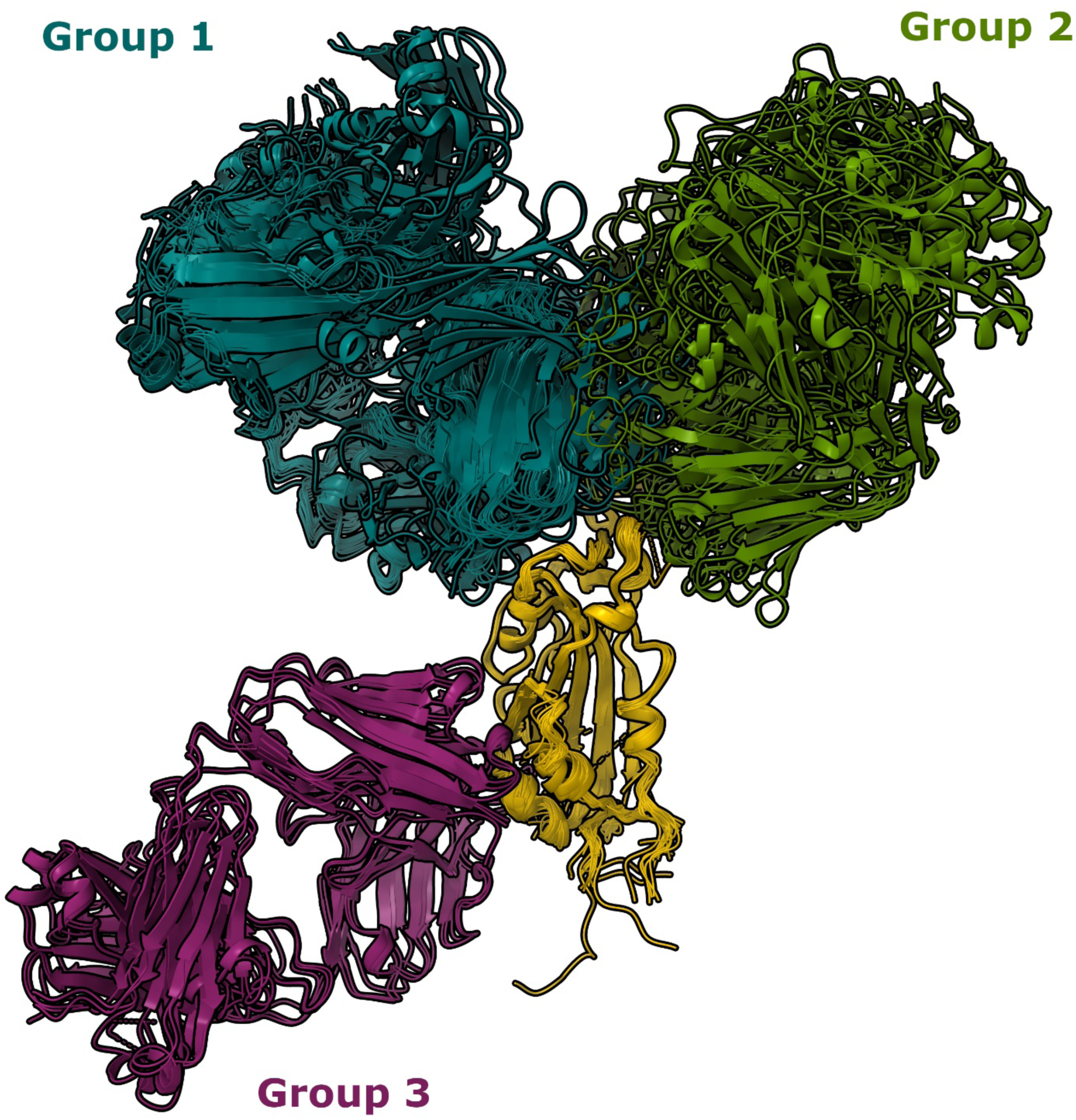
RBD-Ab binding site modes structural comparison. 3D representation of RBD-antibody binding site mode found in the PDB. Structures were superimposed on RBD (yellow cartoons) Cα atoms and colored according to the group they belong to. The first group is composed by twenty structures, the second group includes nine structures and the third 5 structures. Structures with PDBID: 7BEK, 7MF1 and 6YLA were chosen as representatives for each group. See Supplementary Note 1 for more details.

**Extended Data Figure 2.**
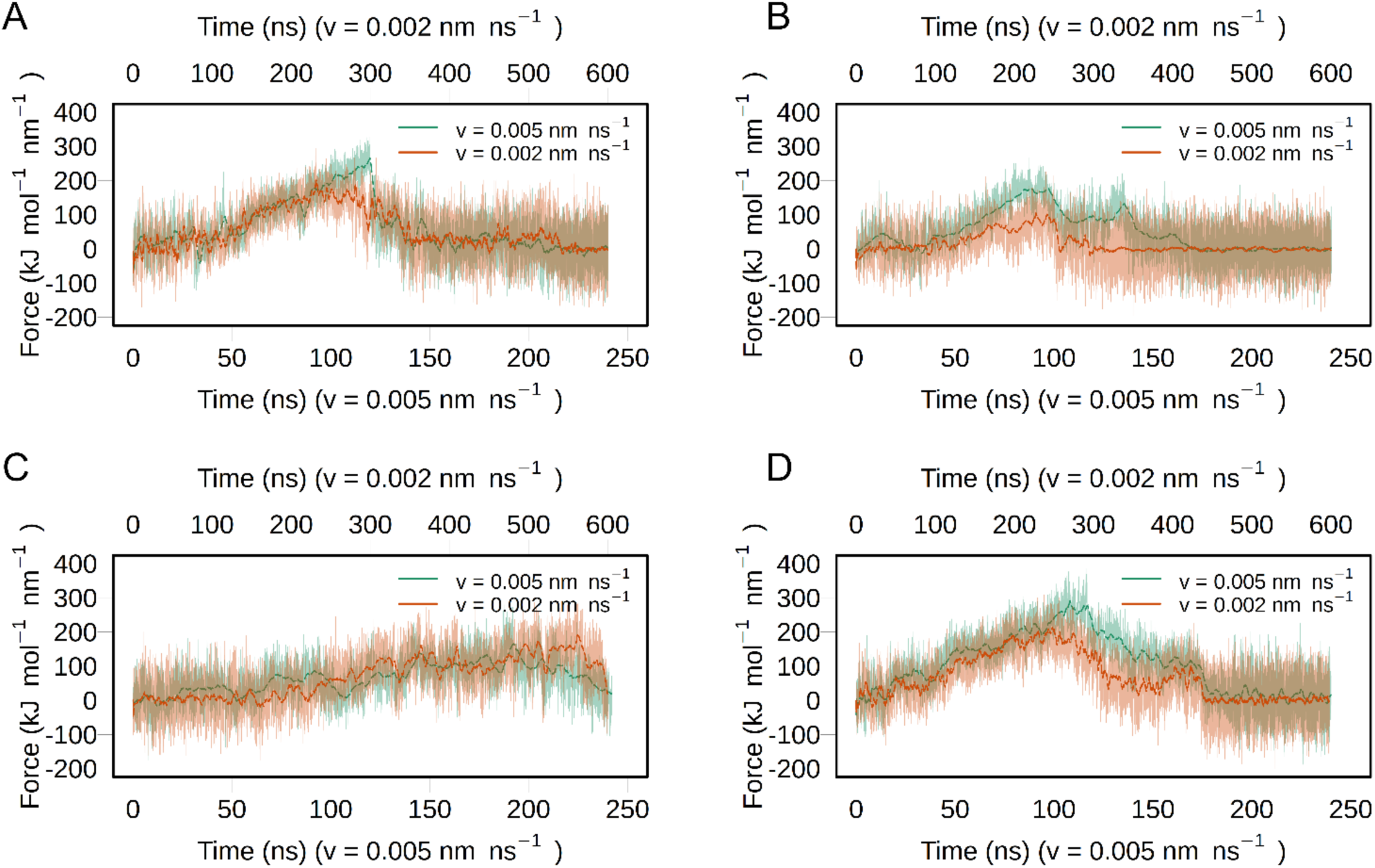
Steered MD force profiles. Force profiles for the SMD simulations of ACE2-RBD (**A**); Ab-RBD from group 1 (**B**); Ab-RBD from group 2 (**C**); Ab-RBD from group 3 (**D**). As expected, simulations at lower pulling speeds (0.02 Å ns^-1^) required lower forces to unbind but present a force-profile similar to simulations at higher speeds. The maximum force observed in simulations at lower pulling speed was around 250 kJ mol^-1^ nm^-1^, for ACE2-RBD and Ab-RBD Group 3, a value higher than those reported in AFM experiments^41^, but expected due to the large difference in the pulling speed used due to computational limitations.

**Extended Data Figure 3.**
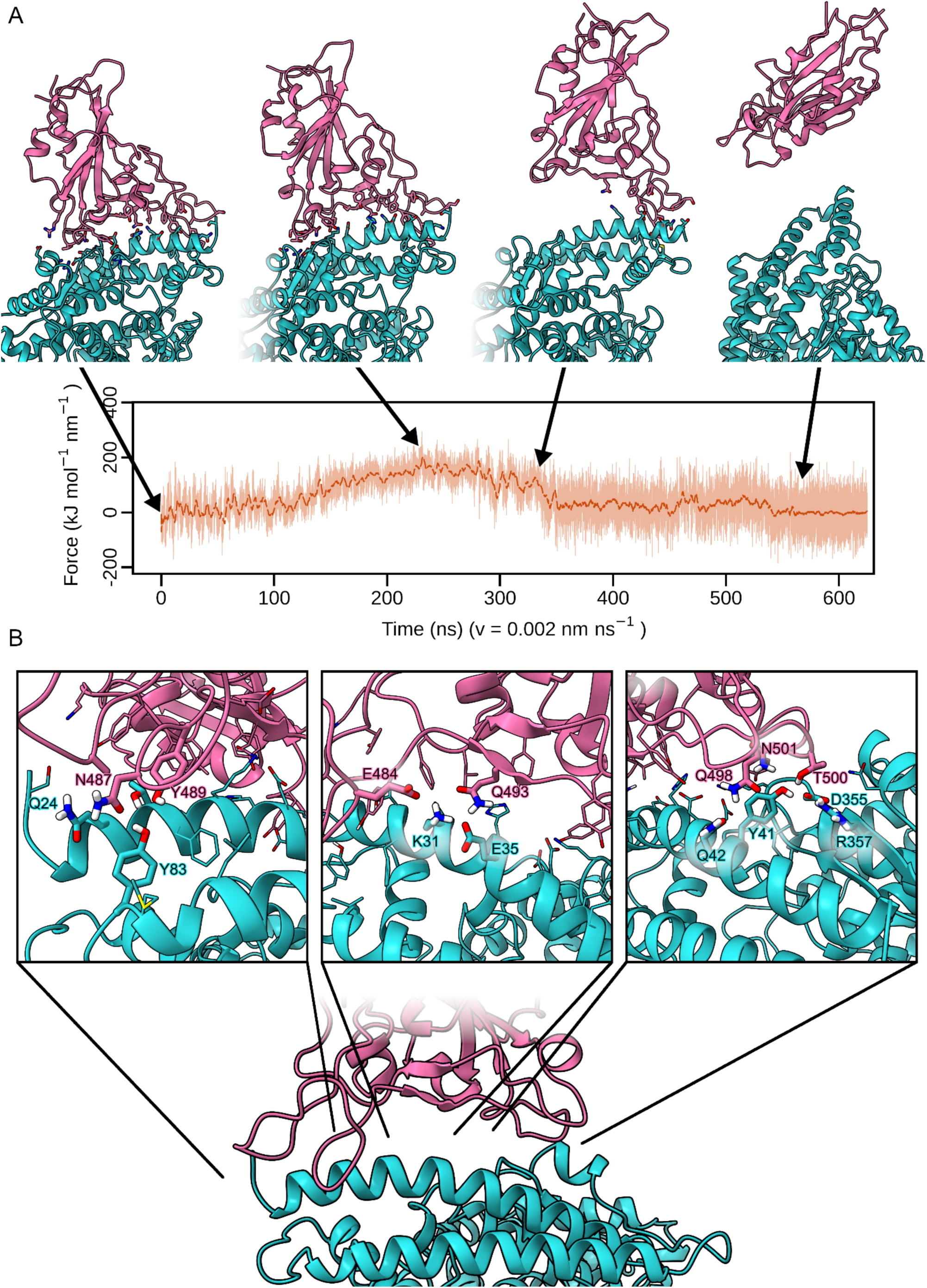
Unbinding dynamics of complexes. RBD unbinding mechanism from ACE2 receptor (**A**) and most stable interactions at the interface (**B**). After the maximum force was reached at about 250 ns, the RBD starts to detach from the ACE2 binding surface but preserving the interactions in the region of the RBD binding ridge (between residues 472-490). Interestingly, the binding conformation of this region presents the most relevant difference compared to the binding mode of the SARS-CoV RBD^42^. From our SMD simulations this region of RBD appears to be the most energetically relevant to maintain a bound conformation with ACE2. In particular, residues N487 and Y489 within the RBD binding ridge were found to be involved in the most persistent interactions with other polar ACE2 residues, such as Q24 and Y83. These interactions were maintained for over 400 ns in the SMD simulation. Other interactions such as residues E484 and Q493 with K31 and E35 or Q498, T500 N501 with Y41, Q42 D355 and R357 were broken after the maximum force was reached at approximately 250 ns.

**Extended Data Figure 4.**
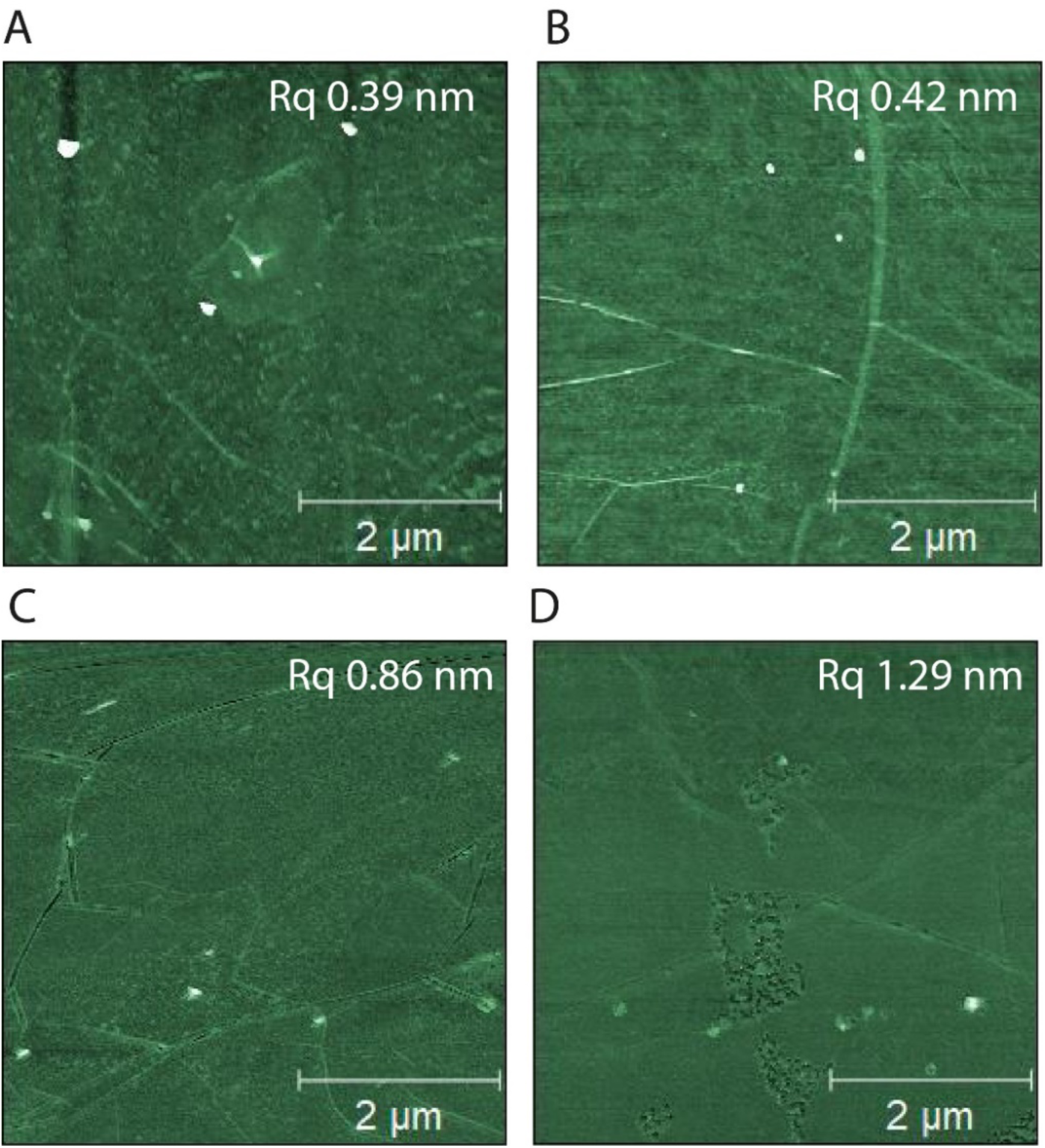
Atomic Force Microscopy images and Rq on PBASE treated graphene. AFM images of untreated gFET (**A**) and after treatment with (**B**) 2 mM, (**C**) 5 mM and (**D**) 10 mM PBASE solution. Rq, root mean surface roughness is reported on each image.

**Extended Data Figure 5.**
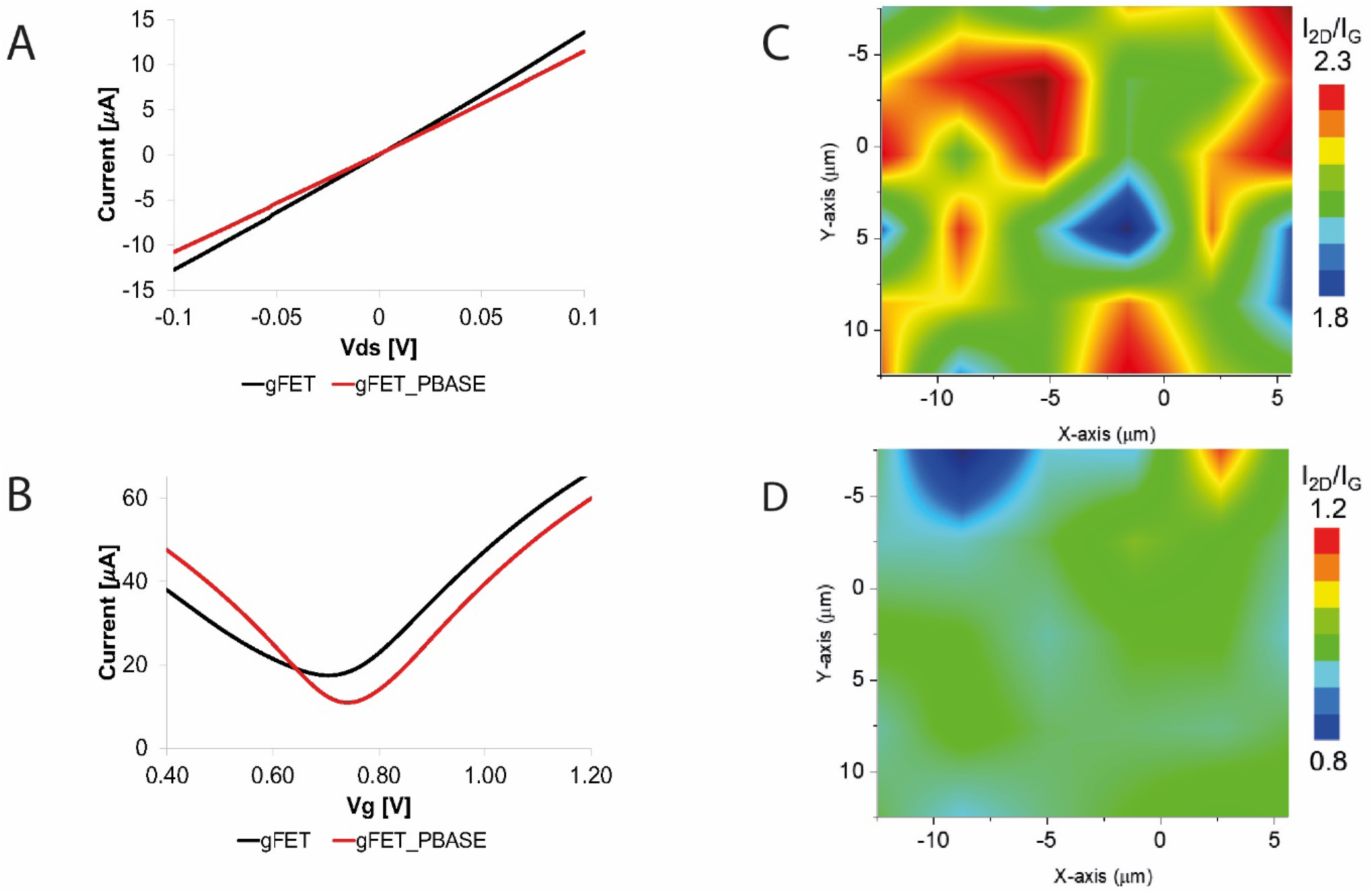
Transfer characteristics and Raman maps on prepared gFET. (**A**) Current-voltage curves (I_ds_ -V_ds_) of gFET (black) and gFET_PBASE (red) obtained *(i)* by applying a V_ds_ between -0.1 V to 0.1 V, *(ii)* by operating in liquid gating condition with PBS solution pH 7.4, and *(iii)* V_g_ 0 V. (**B**) Transfer curves (I_ds_ -V_g_) of pristine gFET (black) and gFET_PBASE (red) obtained *(i)* by using V_ds_ 0.050 V, *(ii)* by operating in liquid gating condition with PBS solution pH 7.4, and *(iii)* by applying V_g_ from 0.4 to 1.2 V. (**C-D**) Raman Spectra maps of gFET in false colour images displaying the distribution of I_2D_ /I_G_ ratio of gFET(**C**) and gFET_PBASE (**D**). Scan size 20 microns.

**Extended Data Figure 6.**
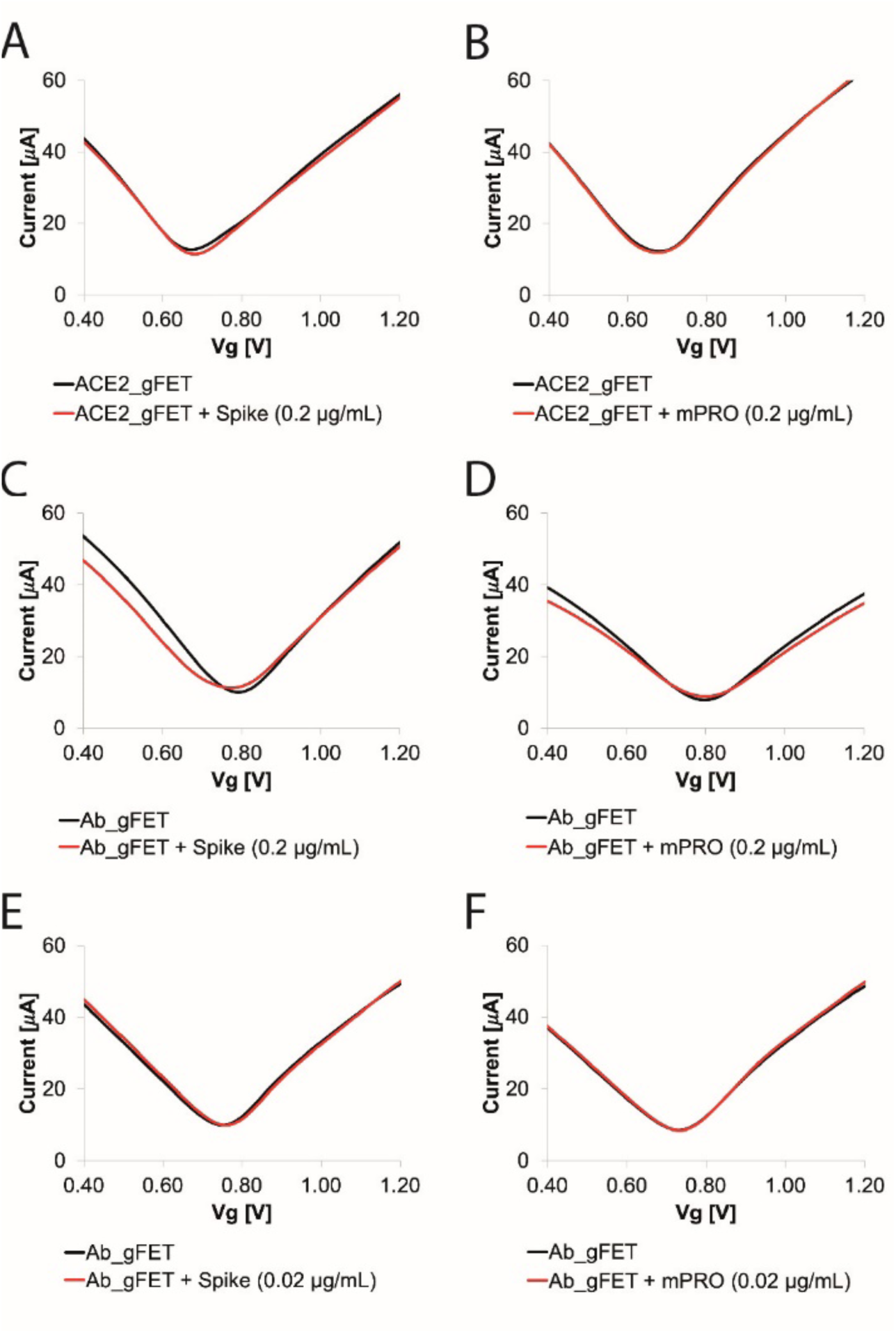
gFET transfer curves in response to Spike and mPRO. Transfer curves for (**A**) ACE2_gFET (black) and ACE2_gFET + Spike (red) at 0.2 µg/mL; (**B**) ACE2_gFET (black) and ACE2_gFET-mPRO + mPRO (red) at 0.2 µg/mL; (**C**) Ab_gFET (black) and Ab_gFET + Spike (red) at 0.2 µg/mL; (**D**) Ab_gFET (black) and Ab_gFET + mPRO (red) at 0.2 µg/mL. (**E**) Ab_gFET (black) and Ab_gFET + Spike (red) at 0.02 µg/mL; (**F**) Ab_gFET (black) and Ab_gFET + mPRO (red) at 0.02 µg/mL. Transfer curves (I_ds_ -V_g_) were obtained *(i)* by using V_ds_ 0.050 V, *(ii)* by operating in liquid gating condition with PBS solution pH 7.4, and *(iii)* by applying V_g_ from 0.4 to 1.2 V.

**Extended Data Figure 7.**
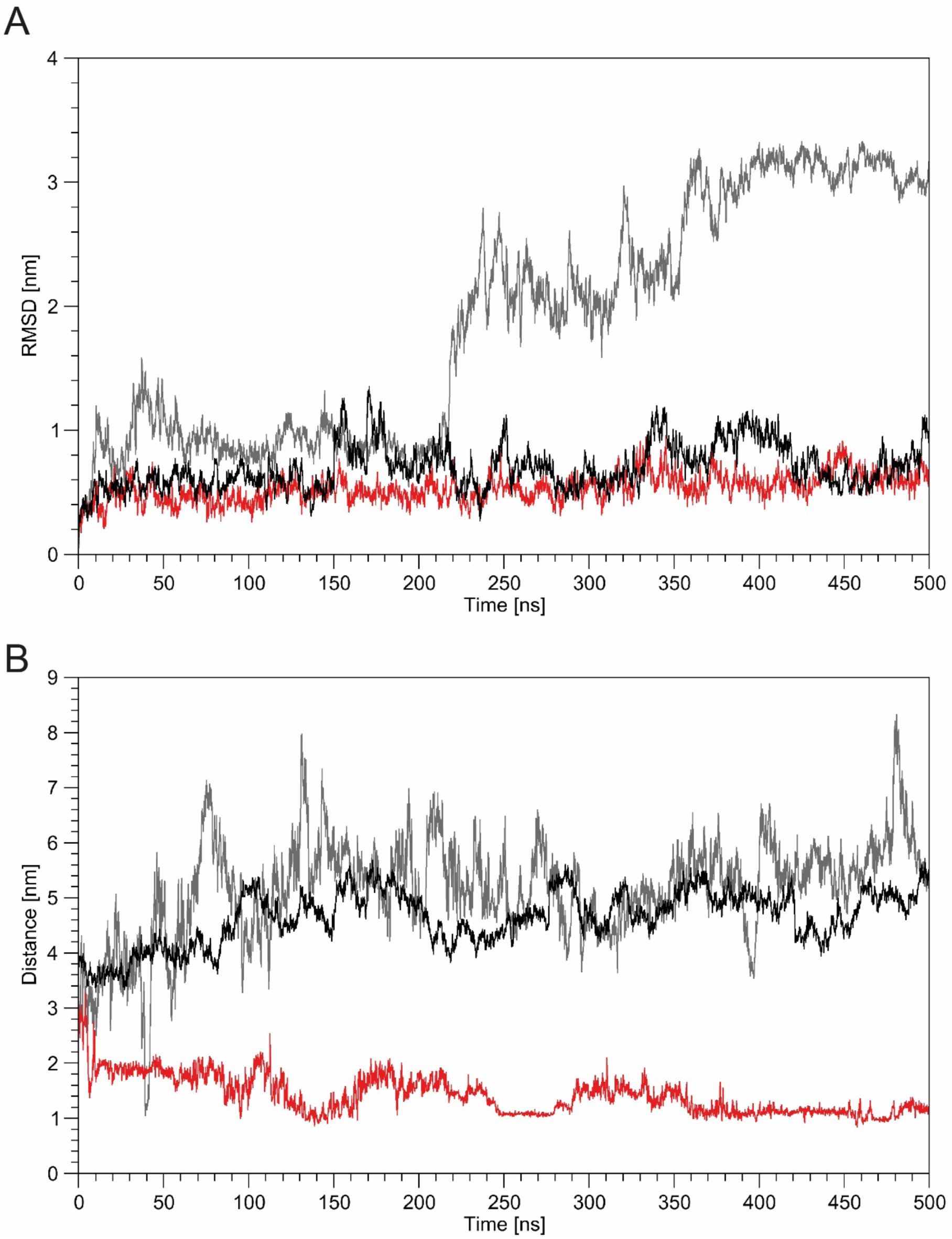
Dynamics of ACE2 dimers in time. (**A**) Time evolution of the Root Mean Square Deviation of the ACE2 PD domains for the ACE2-Membrane (black), ACE2-Fc (red) and soluble ACE2 (grey) systems. (**B**) Time evolution of distance of the CLD-Fc connecting linkers. Distance between the centers of mass of residues lying at the middle of the linker below CLD domain for the ACE2-Membrane (black), ACE2-Fc (red) and soluble ACE2 (grey) systems is shown.

**Extended Data Figure 8.**
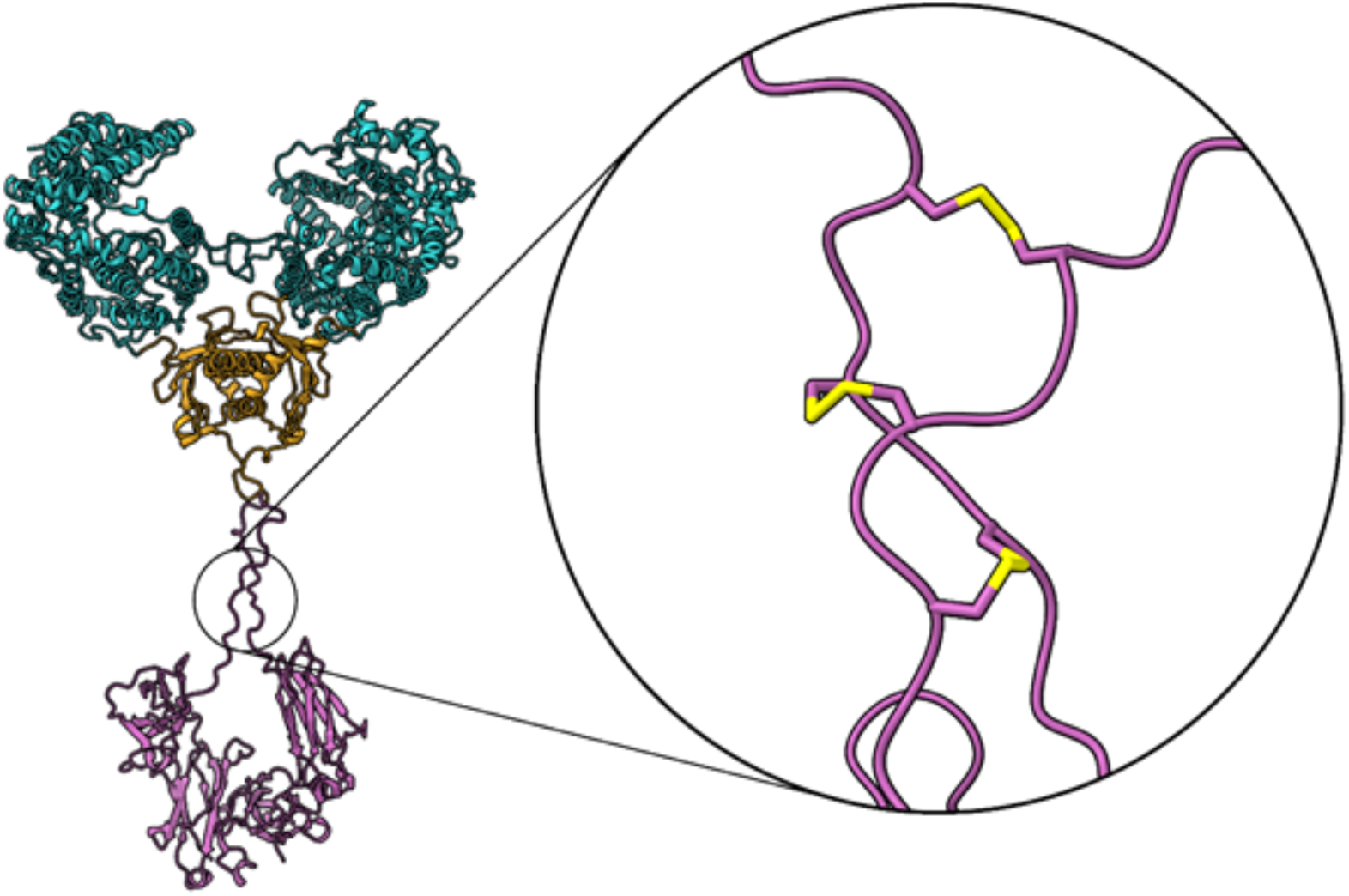
Zoom on the disulfide bridges within the Fc region. Structure of the ACE2-Fc chimera and details of the disulfide bridges connecting monomers

**Extended Data Figure 9.**
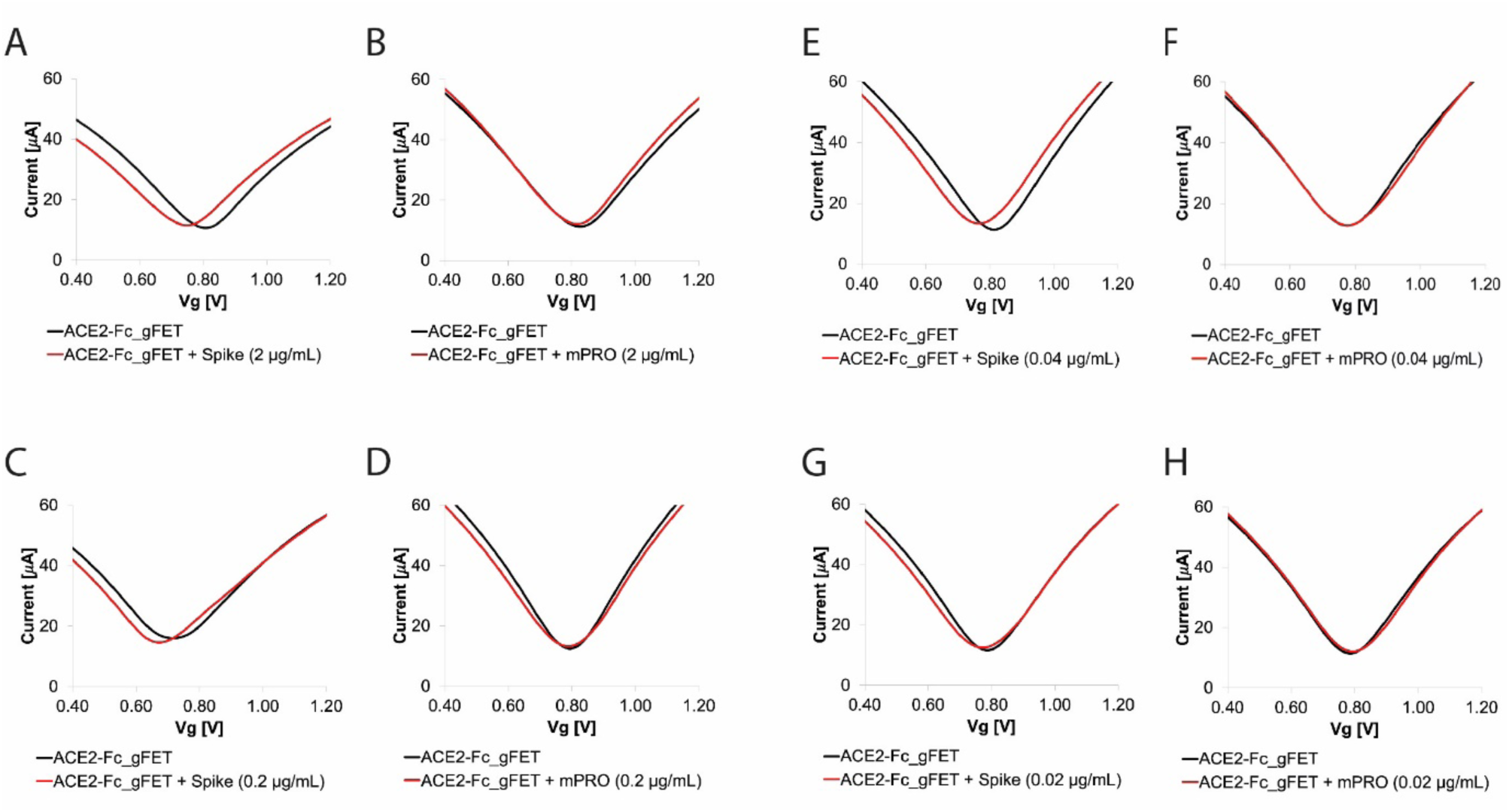
gFET transfer curves in response to Spike and mPRO. Transfer curves for (**A**) ACE2-Fc_gFET (black) and ACE2-Fc_gFET + Spike (red) at 2 µg/mL; (**B**) ACE2-Fc_gFET (black) and ACE2-Fc_gFET + mPRO (red) at 2 µg/mL; (**C**) ACE2-Fc_gFET (black) and ACE2-Fc_gFET + Spike (red) at 0.2 µg/mL; (**D**) ACE2-Fc_gFET (black) and ACE2-Fc_gFET + mPRO (red) at 0.2 µg/mL; (**E**) ACE2-Fc_gFET (black) and ACE2-Fc_gFET + Spike (red) at 0.04 µg/mL; (**F**) ACE2-Fc_gFET (black) and ACE2-Fc_gFET + mPRO (red) at 0.04 µg/mL; (**G**) ACE2-Fc_gFET (black) and ACE2-Fc_gFET + Spike (red) at 0.02 µg/mL; (**H**) ACE2-Fc_gFET (black) and ACE2-Fc_gFET + mPRO (red) at 0.02 µg/mL; Transfer curves (I_ds_ -V_g_) were obtained *(i)* by using V_ds_ 50 mV, *(ii)* by operating in liquid gating condition with PBS solution pH 7.4, and *(iii)* by applying V_g_ from 0.4 to 1.5 V.

**Extended Data Figure 10.**
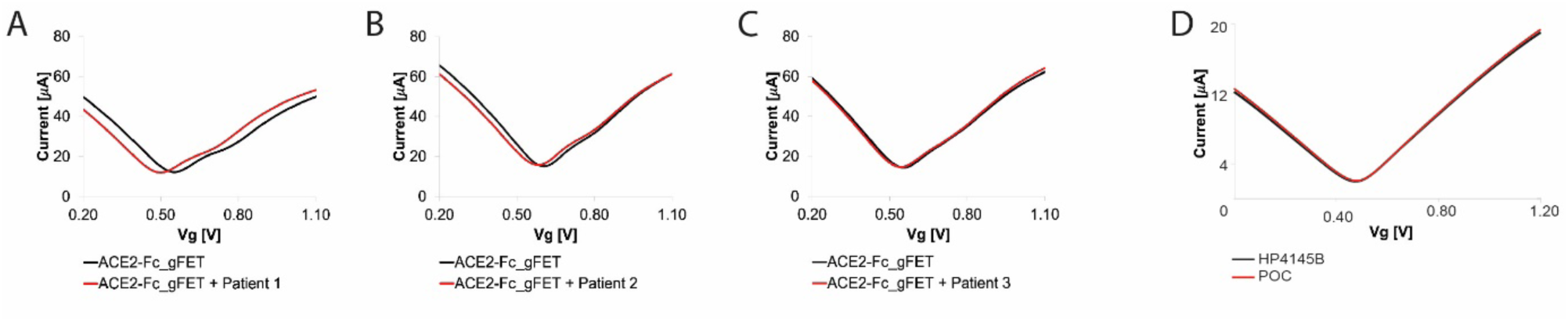
Transfer curves for ACE2-Fc_gFET on patient samples and comparison between curves obtained with POC and semiconductor analyser equipped with probe station. Transfer curves measured by POC for the ACE2-Fc_gFET in response to patient samples: (**A**) Patient 1, (**B**) Patient 2, and (**C**) Patient 3. (**D**) Transfer curves of an unfunctionalized gFET obtained with Semiconductor analyses HP4145B (black) and prototype POC device (red). Transfer curves (I_ds_ -V_g_) were obtained *(i)* by using V_ds_ 50 mV, *(ii)* by operating in liquid gating condition with PBS solution pH 7.4, and *(iii)* by applying V_g_ from 0 to 1.2 V.

**Extended Data Figure 11.**
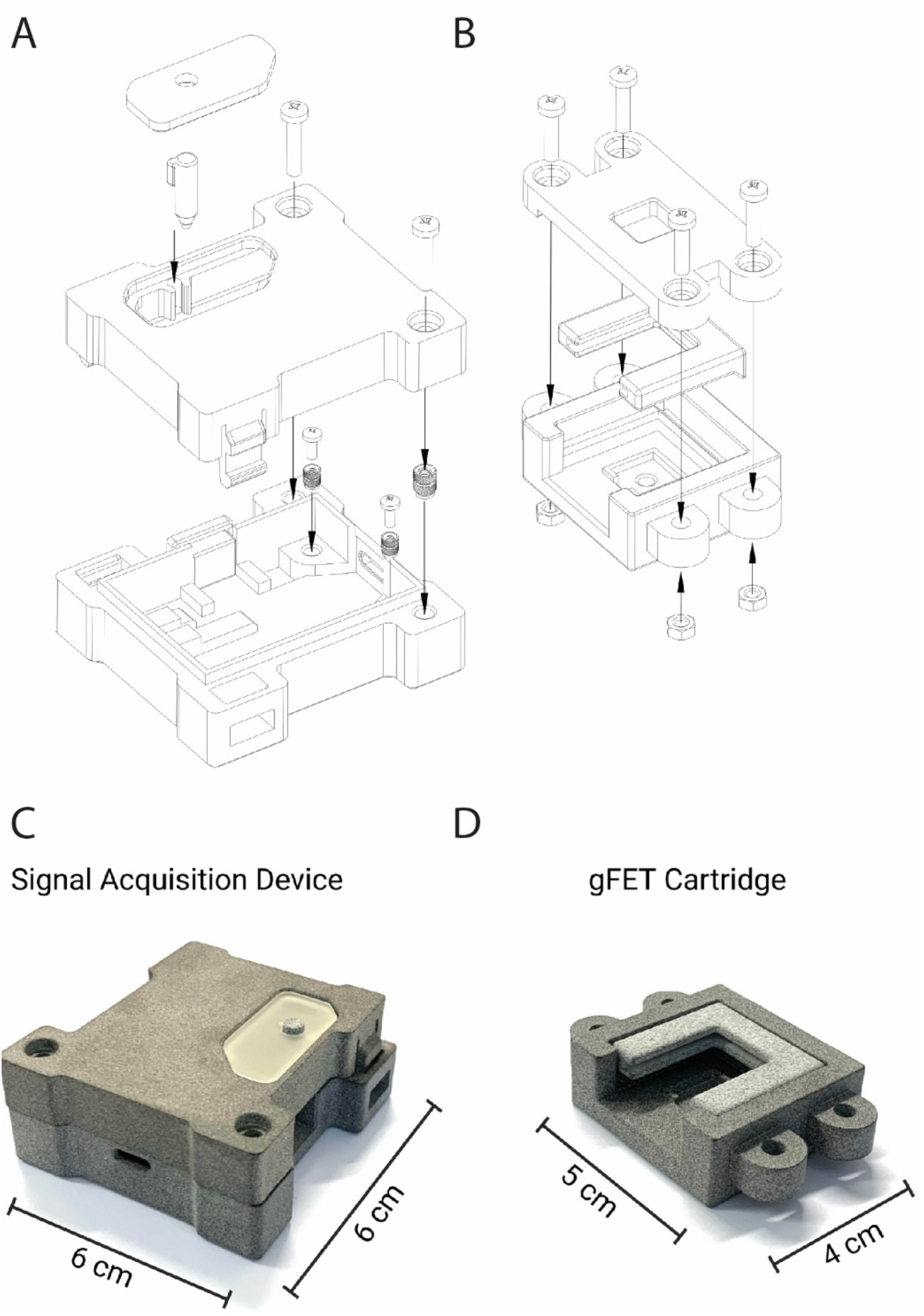
Prototype case and carrier: design and production. (**A**) Case assembly unit: Window, Button, Upper case and Lower case (from top to bottom); (**B**) Carrier assembly unit: Carrier 3, Carrier 1 and Carrier 2 (from top to bottom). Photographs of the device units: main case (**C**) and gFET carrier case (**D**). 3D printed using the SLS (Selective Laser Sintering) printing technique.

**Extended Table 1.** Ab-Spike structures in the PDB. List of the PDB ID codes, resolution and corresponding reference of all the Antibody-Spike complex structures used for the analysis of interaction patterns and structural clustering of binding modes. See Supplementary Note 1 for more details.

| PDB ID | Resolution (Å) | Citation |
| --- | --- | --- |
| 7BEK | 2.04 | (Dejnirattisai et al., 2021)<sup>43</sup> |
| 7MF1 | 2.09 | (Zhou et al., 2021)<sup>44</sup> |
| 7CJF | 2.11 | (Guo et al., 2021)<sup>45</sup> |
| 7BEH | 2.30 | (Dejnirattisai et al., 2021)<sup>43</sup> |
| 7BEI | 2.30 | (Dejnirattisai et al., 2021)<sup>43</sup> |
| 6XC4 | 2.34 | (Yuan et al., 2020)<sup>46</sup> |
| 7CHB | 2.40 | (Du et al., 2020)<sup>47</sup> |
| 7BEJ | 2.42 | (Dejnirattisai et al., 2021)<sup>43</sup> |
| 7KQB | 2.42 | (Asarnow et al., 2021)<sup>48</sup> |
| 6YLA | 2.42 | (Huo et al., 2020)<sup>49</sup> |
| 7BEN | 2.50 | (Dejnirattisai et al., 2021)<sup>43</sup> |
| 7BEM | 2.52 | (Dejnirattisai et al., 2021)<sup>43</sup> |
| 7BEL | 2.53 | (Dejnirattisai et al., 2021)<sup>43</sup> |
| 6XKQ | 2.55 | (Kreye et al., 2020)<sup>50</sup> |
| 7KN6 | 2.55 | (Koenig et al., 2021)<sup>51</sup> |
| 7K43 | 2.60 | (Tortorici et al., 2020)<sup>52</sup> |
| 7BEP | 2.61 | (Dejnirattisai et al., 2021)<sup>43</sup> |
| 7M71 | 2.66 | (Asarnow et al., 2021)<sup>48</sup> |
| 7CHF | 2.67 | (Du et al., 2020)<sup>47</sup> |
| 6XC3 | 2.70 | (Yuan et al., 2020)<sup>46</sup> |
| 7CH5 | 2.70 | (Du et al., 2020)<sup>47</sup> |
| 7CZT | 2.70 | (Yan et al., 2021)<sup>53</sup> |
| 7CHC | 2.71 | (Du et al., 2020)<sup>47</sup> |
| 6XKP | 2.72 | (Kreye et al., 2020)<sup>50</sup> |
| 7KN7 | 2.73 | (Koenig et al., 2021)<sup>51</sup> |
| 7ND9 | 2.80 | (Dejnirattisai et al., 2021)<sup>43</sup> |
| 7CZX | 2.80 | (Yan et al., 2021)<sup>53</sup> |
| 7CZW | 2.80 | (Yan et al., 2021)<sup>53</sup> |
| 7ZCQ | 2.80 | (Yan et al., 2021)<sup>53</sup> |
| 7BWJ | 2.85 | (Ju et al., 2020)<sup>54</sup> |
| 7C01 | 2.88 | (Shi et al., 2020)<sup>55</sup> |
| 6XC7 | 2.88 | (Yuan et al., 2020)<sup>46</sup> |
| 7KQE | 2.88 | (Asarnow et al., 2021)<sup>48</sup> |
| 7M7B | 2.95 | (Asarnow et al., 2021)<sup>48</sup> |

**Extended Table 2.** Members of the clustered structures. List of the members of the individual Clusters found. Where available next to the PDB ID the commercial name for the antibody is indicated in parenthesis.

| Group 1 | Group 2 | Group 3 | Group 4 |
| --- | --- | --- | --- |
| 7BEK | 7MF1 | 6YLA | 7KQE |
| 7CHB | 7CHF (bd-368-2) | 6XC3 (cr3022) | 7M7B |
| 7CHF (bd-604) | 7CHC (bd-368-2) | 6XC7 (CC12.3) |  |
| 6XC3 (CC12.1) | 6XKP | 6XC7 (cr3022) |  |
| 7KN6 (Fab) | 7BWJ |  |  |
| 7BEM | 7ZCX |  |  |
| 7BEI | 7M71 |  |  |
| 7BEJ | 7BEP (COVOX384) |  |  |
| 7CHC (bd-629) | 7KQB |  |  |
| 7CH5 |  |  |  |
| 7CJF |  |  |  |
| 7KN7 (Fab) |  |  |  |
| 6XC4 |  |  |  |
| 7CZQ |  |  |  |
| 7C01 |  |  |  |
| 6XKQ |  |  |  |
| 7BEL (COVOX-88) |  |  |  |
| 7CZT |  |  |  |
| 7ND9 |  |  |  |
| 7BEN (COVOX-253) |  |  |  |

**Extended Table 3:** Multistage equilibration protocol steps. The Simulated dimeric ACE2 systems were subjected to energy minimization with positional restraints, then two heating rounds, and finally to four equilibration steps, gradually lowering the restraints. For each of these steps the table reports the restrains used. Here the Protein BB are the protein backbone heavy atoms; Protein SC are the protein sidechain heavy atoms; Lipid polar head are the O3 cholesterol atom and the P POPC atom; Lipid dihedral are is the torsional angle between C3-C1-C2-O21 and C28-C29-C210-C211 POPC atoms. See Supplementary Note 3 for details.

| Step | Protein BB<br>(kJ mol<sup>-1</sup> nM<sup>-2</sup>) | Protein SC<br>(kJ mol<sup>-1</sup> nM<sup>-2</sup>) | Lipid Polar Head<br>(kJ mol<sup>-1</sup> nM<sup>-2</sup>) | Lipid dihedral<br>(kJ mol<sup>-1</sup> rad<sup>-2</sup>) |
| --- | --- | --- | --- | --- |
| Mini | 4000 | 2000 | 1000 | 1000 |
| Heat 1 | 4000 | 2000 | 1000 | 1000 |
| Heat 2 | 2000 | 1000 | 1000 | 400 |
| Equil 1 | 1000 | 500 | 400 | 200 |
| Equil 2 | 500 | 200 | 200 | 200 |
| Equil 3 | 200 | 50 | 40 | 100 |
| Equil 4 | 50 | 0 | 0 | 0 |

## Supplementary Information File

**Supplementary note 1: Analysis of RBD/antibodies interaction patterns**

In order to obtain a comparison of the putative ACE2 binding sites of the receptor binding domain (RBD) of the Spike protein of SARS-CoV-2, we first retrieved the available structures from the Protein Data Bank website (PDB) and then performed a structural alignment and an evaluation of the Root Mean Square Deviation (RMSD) in order to cluster the found structures in different families. On the PDB website we researched all structure of RBD or Spike protein co-crystallized with human antibodies, with a resolution below 3 Å. All structure belonging to studies not yet published were excluded. We obtained 33 entries, corresponding to 39 RDB-antibodies binary structures (see Extended Table 1). Structures were aligned on the RBD region using the software Chimera^1^. Structures that at visual inspection did not match any other structure (7K43, 7CZW, 7BEP, 7BEN, 7BEL) were excluded from the analysis. Then we calculated the RMSD over residues 10 to 114 of the heavy and light chains of every antibody and organized the structures into four different groups. The first group is composed by twenty structures, the second group includes nine structures, the third five structures and the last group only encompasses two structures (see Extended Table 2).

We then selected a representative structure from each of the three most populated groups to perform simulation aimed at evaluating the forces required to observe the RBD-antibody unbinding events. We chose structure 7BEK for the first cluster, 7MF1 for the second cluster and 6YLA for the third cluster.

### Supplementary note 2: Steered Molecular Dynamics simulations

The structure of ACE2 PD domain bound to SARS-CoV-2 Spike RBD (PDBID: 6VW1^2^) and the three structures representatives of the Ab-RBD binding modes (PDBID: 7BEK^3^, 7MF1^4^ and 6YLA^5^) were prepared using the input generator of CHARMM-GUI^6^. charmm36m forcefield was used in all the simulations^7^. The systems were then pre-aligned maintaining the direction of unbinding parallel to the x axis. Systems were then immersed in a water box with the size of 18 nm × 13.0 nm × 13.0 nm for the ACE2-RBD system, 20 nm × 12.0 nm × 12.0 nm for the 7BEK and 7MF1 systems and 18 nm × 11 nm × 11 nm for 6YLA system. The size of the box was set to accommodate the system and increasing the × dimension of about 4 nm to allow the RBD unbinding. Systems were neutralized with K+ or Cl-counterions according to their total charges. Simulations were run using GROMACS 2018.3^8^. A multistage equilibration protocol, adapted from^9^ was applied to remove unfavourable contacts and provide a reliable starting point for the SMD runs. The systems were first subjected to 1000 step of steepest descent energy minimization with positional restraints (2000 kJ mol^-1^ nM^-2^) on all resolved atoms. Subsequently a 1.0 ns NVT MD simulation was used to heat the system from 0 to 100 K with restraints lowered to 400 kJ mol^-1^ nM^-2^ and then the systems were heated up to 300 K in 2.0 ns during an NPT simulation with further lowered restraint (200 kJ mol^-1^ nM^-2^). Finally, the systems were equilibrated during a NPT simulation for 10 ns with backbone restraints lowered to 50 kJ mol^-1^ nM^-2^. SMD production runs were carried out in NPT using the V-rescale thermostat^10^ (coupling constant of 0.1 ps) while pressure was set to 1 bar with the Parrinello-Rahman barostat^11^ (coupling constant of 2 ps). A time step of 2.0 fs was used, together with the LINCS^12^ algorithm to constrain all the bonds. The particle mesh Ewald method^13^ was used to treat the long-range electrostatic interactions with the cut-off distance set at 12 Å. Short-range repulsive and attractive dispersion interactions were simultaneously described by a Lennard-Jones potential, with a cut off at 12 Å, applying long-range dispersion corrections for energy and pressure.

With structures properly equilibrated, SMD simulations were performed by harmonically restraining the x component of the distance between the centre of mass of the backbone of the two proteins. A force constant of 10 kJ mol^-1^ nM^-2^ was used and the equilibrium value of the distance was changed at a constant velocity (two different simulations were performed for each system at 0.05 Å ns^-1^ and 0.02 Å ns^-1^). The force applied to the harmonic spring was monitored during each simulation.

### Supplementary note 3: Simulation of dimeric ACE2 systems

To compare the dynamic properties of ACE2 embedded in membrane (ACE2-Membrane), ACE2 in solution (ACE2), and the chimeric form of ACE bound to Fc (ACE2-Fc) we built the three systems starting from the 6M17 structure^14^, after the removal of other proteins co-crystallized with ACE2. For the ACE-Membrane system we embedded ACE2 in a POPC:CHOL (90:10) membrane using CHARMM-GUI^6^. For the soluble ACE2 we removed the transmembrane helices (residues 741-768) and added a 6-HIS tag at the C-terminal of each chain. For the third system we built an extended model for the ACE2-Fc linking the extracellular portion of ACE2 (residues 21-740) to the structure of Fc. To this end we ran a BLAST search on PDB with the sequence of the Fc tag provided by the manufacturer (Proteintech, UK) and used the best resulting structure (PDB ID 1IGT). The corresponding Fc fragment was isolated from the entire structure and then attached to ACE2 using Chimera^1^. The chimeric structure was then minimized via Steepest Descent as described in^15^. Solution systems (ACE2 and ACE2-Fc) were equilibrated using the same multistage protocol explained for the SMD simulations. The only difference was that the ACE2-Fc linker were not subjected to positional restraints during the equilibration stage. A different equilibration protocol was used for the ACE2-Membrane system to account for the presence of the membrane that requires a slower equilibration. The system was first subjected to 1000 step of steepest descent energy minimization with positional restraints. Subsequently a 1.0 ns NVT MD simulation was used to heat the system from 0 to 100 K and then the system was heated up to 300 K in 2.0 ns during an NPT simulation. Finally, four equilibration steps of 3 ns, 5 ns, 5 ns and 10 ns respectively were performed gradually lowering the restraints acting on the system. Restraints used in each step and the corresponding atoms on which they are applied are summarized in Extended Table 3.

Production runs for all systems were carried out in NPT using the V-rescale thermostat^10^ (coupling constant of 0.1 ps) was used and pressure was set to 1 bar with the Parrinello-Rahman barostat^11^ (coupling constant of 2 ps). A time step of 2.0 fs was used, together with the LINCS^12^ algorithm to constrain all the bonds. The particle mesh Ewald method^13^ was used to treat the long-range electrostatic interactions with the cut off distance set at 12 Å. Short-range repulsive and attractive dispersion interactions were simultaneously described by a Lennard-Jones potential, with a cut off at 12 Å, applying long-range dispersion corrections for energy and pressure. The first 30 ns of each simulation was discarded as a further equilibration stage, and the subsequent 500 ns were analysed.

### Supplementary note 4: Device prototyping

The POC device is composed of a battery-powered, custom-designed, high-accuracy signal acquisition device (Fig. 5A), connected to a replaceable gFET biosensor cartridge (Fig. 5B). It communicates to a measurement PC through a Bluetooth Low Energy (BLE) wireless connection.

The measuring electronics includes low-leakage input multiplexers (MUX) to allow automatic scan of all the 12 transistors in the cartridge, high-resolution 16-bit DACs to set the gate and drain potentials, buffers to drive the gFET transistors, and an ultra-high-resolution, 31-bit ADC to sense the drain current. The biosensor cartridge is designed to be cheap and easily replaceable, contains no active electronics and only provides electrical connections to the gFET terminals, which are routed to the main device through a 24-pole USB-C connector. This provides enough electrical circuits for all the 12 drain terminals and the gate drive, plus control signals to detect cartridge attachment and connector orientation.

The acquisition device is completely autonomous and can store and then send measured data to a PC by means of a custom protocol developed on top of BLE, designed to be robust against radio interferences and temporary loss of connectivity. Through BLE the measurement parameters can also be altered, if need be, and diagnostic information retrieved. In order to detect very low drain currents, the main design challenge for the circuit was to keep electrical noise low.

To this end, a sophisticated power management (PM) section, based on a Texas Instruments (TI) BQ25155 IC, was designed, and several redundant power domains were defined, so that the power rails for the signal chain could be left as clean as possible of the digital noise. The PMIC takes care of recharging the lithium-polymer battery from a micro-USB charging connector, manages and monitors the main system power rail, produces a clean, regulated +3.0 V analog supply voltage for the DAC and voltage reference, and handles auxiliary power rails which, with custom circuitry, take care of proper power sequencing. A dual ±2.5 V supply is generated by a low-noise post-regulated charge pump (TI LM27762) for the analog chain. The microcontroller (Nordic Semiconductor nRF52840) supervises all system operations and communications and includes its own dedicated power rails.

The operation of the device is basically that of a multiplexed source meter. That is, it provides a voltage to a terminal while simultaneously measuring the supplied current, with great care devoted to controlling leakage currents, since the device is meant to measure with high accuracy currents that can be as low as a few microamperes. Typical transistors cannot be used, as most present a leakage current (due to their internal gate protection diodes) of the same order, rendering the measure meaningless. After a careful selection of the state-of-the art devices we thus chose to use femtoFET transistors (TI CSD15380F3), as they have the lowest specified leakage current of any commercially available MOSFET, less than 25 nA.

These, coupled with a high-input-impedance ADC (TI ADS1284, 1 nA bias current) and op-amps (TI OPA388, 30 pA input bias current), together with precision MUX (TI TMUX1108, 8 pA typical leakage current) still remain the main source of error. To partially overcome these errors, at least the deterministic portion of them, calibration resistors have been inserted in spare MUX channels.

Overall, the circuit provides a 125 µA full-scale readout to the ADC, which, with its 20 bits of effective distortion-free resolution, is able to sense currents well below the aforementioned error sources, making it also possible to operate the gFETs at much lower drain voltages than the 50 mV used in the laboratory experiments, should the need arise.

The DAC that sets the driving voltages (TI DAC80502) has indeed about a 20 µV resolution, and the driving scheme with the MOSFET buffer and bipolar supply of the op-amp allows the system to drive voltages very close to ground, creating a compact, portable system with performances on par to those of a full-fledged rack-mounted lab instrument.

### Supplementary note 5: Designing the Case and Carrier units

The case of the Signal Acquisition Device was based on a 2D CAD drawing of the PCB board. In order to run virtual simulations of the assembly and check for interferences between all parts, a 3D model of the Board, Antennae and Battery was created and ultimately the case ended up being shaped around those objects. In accordance with design requirements passed along by the Engineers who developed and built the PCB, no metal screws could be used in proximity of the antenna in order to avoid possible signal interferences. Furthermore, a 15mm clearance above the PCB was required in order to fit the battery, alongside a plug and cables. Following several iterations, the final design featured a simple rectangular shape, which allowed us to achieve very good results even with traditional 3D printing solutions such as FDM and basic materials like ABS.

The Signal Acquisition Device assembly is comprised by four main parts: an Upper Case, a Lower Case, a Window and a Button (Extended Figure 11). The Upper Case features two holes in the front to host M3 screw heads, two male snap fits in the rear (in proximity of the Antennae) and a slot for fitting a transparent window. The Lower Case acts as a rest bed for the main PCB board and its battery. It features two Ø3,3mm holes at the front (for M2 heat inserts) alongside two Ø4mm holes (for M3 heat inserts). The M2 heat inserts, in combination with M2×5 screws, are used to keep the board in a fixed and stable position. The M3 heat inserts allow (with the use of M3×14 screws) to tightly close the Upper and Lower Case together. The lower case also presents female snap fits at the rear and holes on the sides made to access the USB Type C and the USB Micro B connector. The window allows the user to visually see LED lights (located on the board) while operating the device. It was obtained by pouring transparent epoxy resin into a 3D printed ABS mold. The button acts as an extension and allows to trigger the on/off switch located on the PCB. In order to fully test the snap fit joints, a final prototype was 3D printed using the SLS (Selective Laser Sintering) printing technique (Extended Data Figure 11).

Alongside the main Case, the gFET Cartridge was designed (Extended Data Figure 11). This unit keeps the graphene chip and the spring-loaded contact pins together, while ensuring a stable and precise alignment between the two. It is arguably the most delicate parts of the biosensor, since any small misalignment between these two components would lead to inconsistent or unreliable measurements. The gFET Cartridge is comprised of three main parts: the Carrier 1, Carrier 2 and Carrier 3 (Extended Data Figure 11). The Carrier 1 houses the cartridge where the spring-loaded contact pins are encapsulated. The Carrier 2 has a slot that contains the 10.4×10.4mm gFET chip. Early prototypes featured larger slots to take into account tolerances and the 3D printer’s accuracy, however, after confirming that a high printing quality could be achieved with a traditional FDM printer, clearance was decreased by narrowing down the gFET housing slot. The result was a much higher measuring precision and reliability. The Carrier 2 also features hexagonal-shaped housings into which M3 nuts can be snapped into place. The Carrier 3 acts as a plate that holds the whole assembly together with the help of M3×12 screws. An opening allows a lab pipette to access and reach the gFET inside. Despite being part of the assembly design, final tests showed that the Carriers 1 and 2 had such a precise fit and low clearance that the device could be operated even without the Carrier 3 part installed.

**Supplementary Video 1. Comparison of MD simulations of dimeric ACE2 in our three systems**. Left: full-length immersed in a lipidic membrane; middle: chimeric ACE2-Fc in water; right: soluble ACE2 in water.

